# First-in-human, phase 1, randomized, observer-blind, controlled trial to assess the safety and immunogenicity of novel live attenuated type 1 and type 3 oral poliomyelitis vaccines in healthy adults

**DOI:** 10.1101/2025.02.21.25322566

**Authors:** Laina D. Mercer, Arlene C. Seña, E. Ross Colgate, Jessica W. Crothers, Peter F. Wright, Mohamed Al-Ibrahim, Erman Tritama, Annelet Vincent, Bernardo A. Mainou, Yiting Zhang, Jennifer Konopka-Anstadt, Ananda S. Bandyopadhyay, Alan Fix, John O. Konz, Chris Gast

## Abstract

**Background:** Reducing the risks of vaccine-derived polioviruses and vaccine-associated paralytic poliomyelitis from type 1 or 3 Sabin-strain oral poliovirus vaccines (OPVs) motivated the development of novel type 1 and 3 OPVs (nOPV1, nOPV3), designed to have similar safety and immunogenicity and improved genetic stability to reduce risk of reversion to neurovirulence. In this first-in-human trial, we assessed safety and immunogenicity of nOPV1 and nOPV3 in healthy adults.

**Methods:** We conducted a multi-site, randomized, observer-blind, controlled trial in healthy adults in the United States. Participants were stratified according to poliovirus vaccination history (exclusive inactivated polio vaccine [IPV] or including OPV) and randomized to receive either nOPV or homotypic Sabin-strain monovalent OPV (mOPV); IPV participants received a single dose and OPV participants received two doses. The primary objective was to assess safety measured by adverse events. The secondary objectives were to assess serum neutralizing antibody responses measured before and 28 days after each dose and fecal viral shedding assessed up to 56 days post-first dose. This study was registered with ClinicalTrials.gov, NCT04529538.

**Findings:** Between May 2021 and February 2023, 205 healthy adults were enrolled and received at least one dose: 70 nOPV1, 45 mOPV1, 56 nOPV3, and 38 mOPV3. Most events were mild, severe events were rare, and solicited events were balanced. Homotypic seroprotection was nearly 100% at baseline and was 100% after the first dose. Homotypic seroconversion rates after a single dose were high and similar for nOPV and mOPV (from 86 to 100%), with no statistically significant differences. Similar rates of viral shedding were observed among participants receiving nOPV or mOPV.

**Interpretation:** Both nOPV1 and nOPV3 were well tolerated and demonstrated similar immunogenicity and shedding profiles to mOPV1 and mOPV3, respectively, supporting progression to phase 2 studies. nOPVs may be an important tool for achieving eradication of poliovirus.

**Funding:** Gates Foundation.

**Research in Context:** *Evidence before this study:* Sabin-strain vaccine-derived polio virus (cVDPVs) and vaccine-associated paralytic polio (VAPP) are now a substantial proportion of paralytic poliomyelitis worldwide. To reduce the seeding of type 2 cVDPVs (cVDPV2), a more genetically stable novel oral polio vaccine (nOPV2) was developed to control outbreaks. WHO granted use under emergency use listing (EUL) in 2020 and prequalified the vaccine in 2023. More than one billion doses have been distributed since March 2021, with surveillance data demonstrating a promising safety and effectiveness profile. Sabin-strain types 1 and 3 present similar risks for cVDPVs and VAPP. In pre-clinical studies chimeric viruses with nOPV2’s non-structural regions, including changes to the RNA sequence in the 5’ untranslated region, the non-structural protein 2C, and the polymerase 3D, coupled with the coding region for the type-specific Sabin-strain capsid proteins have demonstrated similar immunogenicity, antigenicity, and lower neurovirulence compared to Sabin.

*Added value of this study:* This first-in-human trial includes safety and immunogenicity data in adults with a history of either exclusive inactivated polio vaccine (IPV) or prior exposure to OPV. We found that nOPV1 and nOPV3 are safe, well tolerated, and induce similar immunogenicity to their Sabin controls. The magnitude and durations of nOPV shedding was not higher than Sabin controls. We also observed induction of mucosal immunity, evidenced by reduced viral shedding post second vaccination.

*Implications of all the available evidence:* The successful deployment of nOPV2 to combat cVDPV2s previously demonstrated that use of such novel vaccines can be effective in the control of cVDPV outbreaks after the cessation of Sabin-strain types 1 and 3. nOPVs can thus support the polio endgame strategy by providing outbreak response vaccines less likely to be associated with VAPP and seeding of new cVDPVs. The safety and immunogenicity evidence generated for nOPV1 and nOPV3 in this phase 1 clinical study were sufficiently strong to justify phase 2 studies in geographically relevant target populations of previously vaccinated children and infants, as well as vaccine naïve neonates.

## Introduction

Since the Global Polio Eradication Initiative was established in 1988, the use of Sabin-strain oral poliovirus vaccines (OPVs) for immunization has dramatically reduced the burden and transmission of poliomyelitis and was a major contribution to the eradication of wild types 2 and 3 poliovirus. (1) These vaccines provide individual protection and interrupt person-to-person transmission of polioviruses. However, on extremely rare occasions, use of OPV can result in vaccine-associated paralytic polio (VAPP) or paralytic outbreaks due to circulating vaccine-derived poliovirus (cVDPV). (2) Central to both VAPP and cVDPV-induced disease is reversion of the vaccine strain to a more neurovirulent phenotype during intestinal replication of the virus. In case of cVDPVs, reverted viruses can be transmitted to household or community contacts and cause disease outbreaks in settings of persistently poor immunization coverage.

Due to the risks of VAPP and cVDPVs, the withdrawal of routine and supplementary immunization activities with all OPVs, after the eradication of wild polioviruses (WPVs), will be the final stage of polio eradication. (2) The certification of eradication of wild type 2 in 2015, and the predominance of type 2 among cVDPVs, led to a coordinated global discontinuation of the routine use of Sabin-strain type 2 (OPV2) in April 2016. However, as the cohort of children without intestinal mucosal immunity against poliovirus type 2 grew following type 2 OPV cessation, and as IPV coverage remained insufficient, suboptimal vaccination response with monovalent Sabin-strain OPV2 (mOPV2) campaigns against on-going cVDPV2 outbreaks seeded new type 2 cVDPV (cVDPV2) outbreaks. (3), (4)

In parallel, novel type 2 OPV (nOPV2) was developed and introduced to reduce the risk of cVDPV2 outbreaks and VAPP cases. nOPV2 includes modifications to the Sabin 2 genome to stabilize domain V (the primary determinant of attenuation), and includes relocation of the cis-acting replication element and modifications to the polymerase to reduce recombination risk and enhance fidelity of replication. Phase 1 and 2 clinical trials demonstrated nOPV2 to be safe, immunogenic, and more genetically stable than the Sabin-strain OPV2. (5–7) Based on these results, nOPV2 was deployed for outbreak control under World Health Organization (WHO) Emergency Use Listing (EUL), granted in November 2020 (8). Following completion of clinical development (9, 10), nOPV2 was prequalified by WHO in December 2023. (11) More than one billion doses of nOPV2 have now been deployed, and surveillance data are promising in terms of effectiveness and safety. (11–13) The estimated emergence risk of cVDPV2 has been substantially reduced, although not completely eliminated. (14)

Sabin-strain OPV1 and OPV3, like Sabin-strain OPV2, pose rare but potentially consequential risks for VAPP and VDPVs, necessitating development of genetically stable alternatives. Candidate viruses for both polioviruses type 1 and type 3 (nOPV1 and nOPV3, respectively) are chimeric viruses with nOPV2’s non-structural regions coupled with Sabin-strain type 1 or type 3 structural proteins. Compared to the Sabin-strain OPVs, these novel candidates have similar pre-clinical immunogenicity and antigenicity, markedly lower neurovirulence in a transgenic mouse disease model, and reduced reversion to neurovirulence during cell culture passaging. (15) The current trial is the first-in-human evaluation of nOPV1 and nOPV3 in healthy 18–45-year-old adults.

## Methods

### Study Design

We conducted a first-in-human, observer-blind, multi-center randomized controlled study of the safety and immunogenicity of nOPV1 and nOPV3 at University of Vermont Vaccine Testing Center (Burlington, VT, USA), University of North Carolina Institute for Global Health and Infectious Diseases (Chapel Hill, NC, USA), Dartmouth-Hitchcock Medical Center (Lebanon, NH, USA), and Pharmaron (Baltimore, MD, USA). The protocol was approved by Advarrra Institutional Review Board (IRB), University of Vermont IRB, University of North Carolina Biomedical IRB, and Dartmouth-Hitchcock Health Human Research Protection Program. Testing of serum and stool samples was reviewed by the Centers for Disease Prevention and Control (CDC), and was conducted consistent with applicable federal law and CDC policy (See e.g., 45 C.F.R. part 46; 21 C.F.R. part 56; 42 U.S.C. §241(d), 5 U.S.C. §552a, 44 U.S.C. §3501 et seq.)

### Participants

Participants were aged 18-45 years at enrollment and previously received either 3 or more doses of IPV and no OPV (cohorts 1 and 3; referred to as IPV participants) or a full primary polio immunization series containing OPV (cohorts 2 and 4; referred to as OPV participants). Prior vaccination status was ascertained through a combination of vaccination records, self-report, and participant age relative to OPV/IPV switch dates. After providing written informed consent, the serum of potentially eligible participants was screened to confirm neutralizing antibody titre ≥1:8 (seropositivity) for poliovirus type 1 (for cohorts 1 and 2) or type 3 (for cohorts 3 and 4), absence of clinically significant medical conditions, and, if female, a negative serum pregnancy test. Participants were required to reside in the study area and not travel outside the United States until confirmation of cessation of vaccine virus shedding. Females could not be breastfeeding nor pregnant, and those of child-bearing potential agreed to practice adequate contraception for 30 days prior to the first study vaccination and for at least 90 days following the last study vaccination. Participants were excluded for conditions that might be adversely affected by participation, medical conditions that might affect immune responses, professional food handling, residing in a home with a septic tank, and close contact with persons who were immunosuppressed, pregnant, or who had not completed their primary polio immunization series.

### Randomization and masking

Cohorts receiving type 1 vaccines were prioritized for enrollment to prioritize development, given the greater concern about WPV1 and cVDPV1 than cVDPV3. Forty IPV participants were randomized 1:1 to receive one dose of nOPV1 or monovalent Sabin-strain OPV1 (mOPV1) (cohort 1), while 75 OPV participants were randomized 2:1 to receive two doses of nOPV1 or mOPV1 (cohort 2). After cohort 1 was fully enrolled, 36 additional IPV participants were randomized 1:1 to receive one dose of nOPV3 or monovalent Sabin-strain OPV3 (mOPV3) (cohort 3). Similarly, after cohort 2 was fully enrolled, 54 additional OPV participants were randomized 2:1 to receive two doses of nOPV3 or mOPV3 (cohort 4). The sample sizes of cohorts 3 and 4 were lower than those for 1 and 2 due to enrollment challenges and recognition that these sample sizes would still support study objectives. IPV participants received one dose to permit viral shedding observation for two months post-vaccination, while OPV participants provided safety data following each of two doses, prior to multidose trials in younger populations.

Block randomization, stratified by study site, ensured balance within cohorts within each site, without a prespecified number enrolled at each site. The randomization was generated and maintained by the Emmes Company, LLC. Participants withdrawn before vaccination were replaced. The site pharmacists, with primary responsibility for dispensing study vaccines, were charged with maintaining security of the treatment assignments.

### Procedures

Both control vaccines were Sabin-strain mOPVs which are components of the WHO prequalified bivalent OPV (bOPV) manufactured by Bio Farma (Indonesia). The mOPV1 control vaccine contained ≥10^6·0^ 50% cell culture infective dose (CCID_50_) per 0·1mL dose (lot number 2180120), and the mOPV3 control vaccine contained ≥10^5·8^ CCID_50_ per 0·1mL dose (lot number 2080120), the standard dose-levels in bOPV.

Both nOPVs are live, attenuated polioviruses derived from a modified Sabin-strain type 2 infectious cDNA clone and propagated in Vero cells. (16) Nonstructural modifications based on nOPV2 were introduced in the viral nucleotide sequences in the 5’ untranslated region (UTR) to improve and stabilize attenuation, with an element from the 2C coding region relocated to the 5’ UTR to safeguard loss of these modifications through single recombination. Additional modifications to the 3D polymerase reduce the frequency of mutation and recombination. Finally, the type 2 capsid-coding region of the genome was replaced with the capsids from Sabin-strain type 1 (nOPV1) or Sabin-strain type 3 (nOPV3) clones, resulting in chimeric viruses with nOPV2 non-structural regions and Sabin type 1 or 3 structural proteins. The nOPVs contained 10^6·5^ ^± 0·5^ CCID_50_ per 0·1mL dose (lots 2110120 and 2310120 for nOPV1 and nOPV3, respectively), reflecting the highest dose anticipated to be administered in a phase 2 study.

IPV participants in cohorts 1 and 3 received a single dose of vaccine on day 1. OPV participants in cohorts 2 and 4 received a dose on Day 1 and on Day 29. Safety events were reported from the time of informed consent signature through completion of the study 168 days after initial study vaccination (Day 1). Each participant was observed for at least 30 minutes after vaccine administration for immediate adverse reactions. Solicited adverse event (AE) data were collected for seven days (day of study vaccination and six following days) after each dose of study vaccine. If a solicited AE continued beyond seven days, it was reported solely as a solicited AE. Unsolicited AE data were collected for 28 days (day of study vaccination and 27 following days) after each dose of study vaccine. Serious adverse event (SAE) data were collected from the initial study vaccination through the end of the study participation (Day 169).

Clinical laboratory assessments (complete blood count and blood chemistry) were conducted on Days 1 and 8. Blood samples to evaluate vaccine safety were obtained and processed at the trial sites and transported to each sites’ designated laboratory for testing. The severity of any abnormal clinical safety laboratory test results reported as AEs were graded based on the US Food and Drug Administration toxicity grading scales for healthy volunteers in vaccine trials. (17)

For the assessment of humoral immunogenicity, serum samples were collected at the screening visit and on Day 29, and on Day 57 for cohorts 2 and 4. All samples were aliquoted and stored at ≤-20°C before shipping to the Polio and Picornavirus Branch at the United States CDC to measure type-specific polio neutralizing antibodies. (18) Seroprotection was defined as neutralizing antibody titre ≥1:8 for poliovirus. To establish eligibility (seropositivity for the serotype to be administered) prior to enrollment, an aliquot of the screening sample was evaluated in a similar assay at Quest Diagnostics.

Stool samples were collected on Days 1, 8, 15, 22, 29, 36, 43, 50, and 57 for all cohorts and additionally on Days 3, 5, and 10 for cohorts 1 and 3 to detect and assess the quantity of vaccine virus shed, genetic stability of shed polioviruses, and to confirm cessation of shedding. Samples were processed at the trial sites and stored at ≤-20°C before being transported to the CDC or Cerba Research (Rotterdam, The Netherlands; formerly Viroclinics Biosciences B.V.). The detection of type-specific poliovirus in stool was determined via multiplex real-time polymerase chain reaction (PCR) at the CDC. In samples positive solely for type 1 in cohorts 1 and 2 or solely type 3 in cohorts 3 and 4, infectious virus was quantified as the CCID_50_ per gram of stool at the CDC. Next-generation sequencing (NGS) and neurovirulence of shed virus, as assessed by a transgenic mouse neurovirulence test were performed by Cerba; results of which will be reported elsewhere.

### Outcomes

The primary objective was to evaluate the safety and tolerability of nOPV1 and nOPV3 relative to active controls. Primary safety outcomes included the frequency of solicited AEs for seven days after each dose, the frequency of unsolicited AEs for 28 days after each dose (including clinically significant laboratory values on Day 8 reported as AEs), and SAEs from Day 1 through study completion.

Secondary objectives included assessments of serum neutralizing antibody (NAbs) titres compared to type-specific Sabin controls. Type-specific NAbs measured at baseline and 28 days post-vaccination were summarized by median and the geometric mean titres (GMTs). Additionally, post-vaccination type-specific baseline-adjusted GMT ratios and seroprotection (titre ≥1:8) and seroconversion, were assessed. Seroconversion was defined as a ≥4-fold rise in titre in those with titres above the lower limit of quantification (LLOQ; 2·5 log_2_) at baseline or ≥4-fold above LLOQ (4·5 log_2_) for those with baseline titres at or below the LLOQ. Seroconversion was only assessed for participants with baseline titres at least 2·0 log_2_ below the upper limit of quantification (ULOQ; 10·5 log_2_), such that seroconversion could be observed.

Additional secondary objectives included assessment of fecal viral shedding in IPV participants (cohorts 1 and 3); the proportion of participants positive for poliovirus at each stool collection via PCR and via cell culture; the amount of vaccine virus in each stool sample (log_10_ CCID_50_ per gram), averaged over 7-, 14-, 21-, and 28-days post vaccination (shedding index); and the area under the viral shedding curve were calculated. Additionally, time (days) to cessation of fecal shedding, defined as 2 consecutive PCR-negative specimens obtained ≥24 hours apart, was assessed in all participants.

Exploratory outcomes included additional viral shedding and immunogenicity assessments. Heterotypic immune responses (to viral serotypes other than that of the administered vaccine) were also assessed. Shedding outcomes included assessment in OPV participants (cohorts 2 and 4) of all secondary outcomes assessed for IPV participants (cohorts 1 and 3), as well as NGS and mouse neurovirulence in select stool samples from IPV participants. Genetic stability data are outside the scope of this publication and will be reported elsewhere.

### Statistical analysis

The sample sizes for this study were primarily chosen to enable evaluation of safety in a sufficient number of adults to support a subsequent study in younger participants. With at least 50 participants receiving nOPV there was a 95% probability of observing at least one AE if the rate was ≥5.8%, and the upper bound of the 95% confidence interval (CI) with no events observed would be 7.1%. Immunogenicity was a secondary objective and primarily descriptive in nature. Comparisons to the relevant controls were planned, but the sample size was not based on achieving a specific level of statistical power for these comparisons. Focusing on the OPV participants receiving nOPV, with at least 48 (cohort 2 assumption) or 28 (cohort 4 assumption) participants receiving nOPV, if a true seroconversion rate was 50% (maximum variability), the CI for the nOPV response rate would be expected to average ±15% or ±20%, respectively.

Dichotomous outcomes, including safety, seroconversion, and vaccine virus shedding positivity were summarized by counts, proportions, and exact 95% CIs. The median and corresponding bootstrap-based 95% CI were calculated for log_2_ NAbs and for log_10_ CCID_50_/g of shed virus. GMTs were estimated with likelihood-based methods to accommodate censoring at the assay LLOQ and ULOQ. GMT ratios were estimated via a linear model of log_2_ NAb titre as a function of group, site, and baseline log_2_ NAb and similarly accommodated censoring. Two-sided 95% CIs were computed by back-transformation of the CIs derived from the model.

Safety and shedding outcomes were evaluated for all participants who received at least one dose of study vaccine. The per-protocol population (PPP) for immunogenicity analyses was defined as all participants who correctly received study vaccinations per randomization with no major protocol deviations determined to potentially interfere with the immunogenicity result of the participant. Deviations were reviewed and exclusions from the PPP (at the time point level) were identified prior to unblinding.

## Results

The first participant was enrolled on 6 May 2021, and the last participant follow-up was on 17 February 2023. Three hundred and seventy-seven volunteers were assessed for eligibility and 205 were randomized and vaccinated. Major reasons for ineligibility included inability to comply with study restrictions and procedures, presence of chronic or acute health conditions including out-of-range screening laboratory values, or close contact with immunosuppressed or incompletely vaccinated individuals. Seventy participants received at least one dose of nOPV1, 45 received at least one dose of mOPV1, 54 received at least one dose of nOPV3, and 36 received at least one dose of mOPV3 (figure 1).

**Figure 1.**
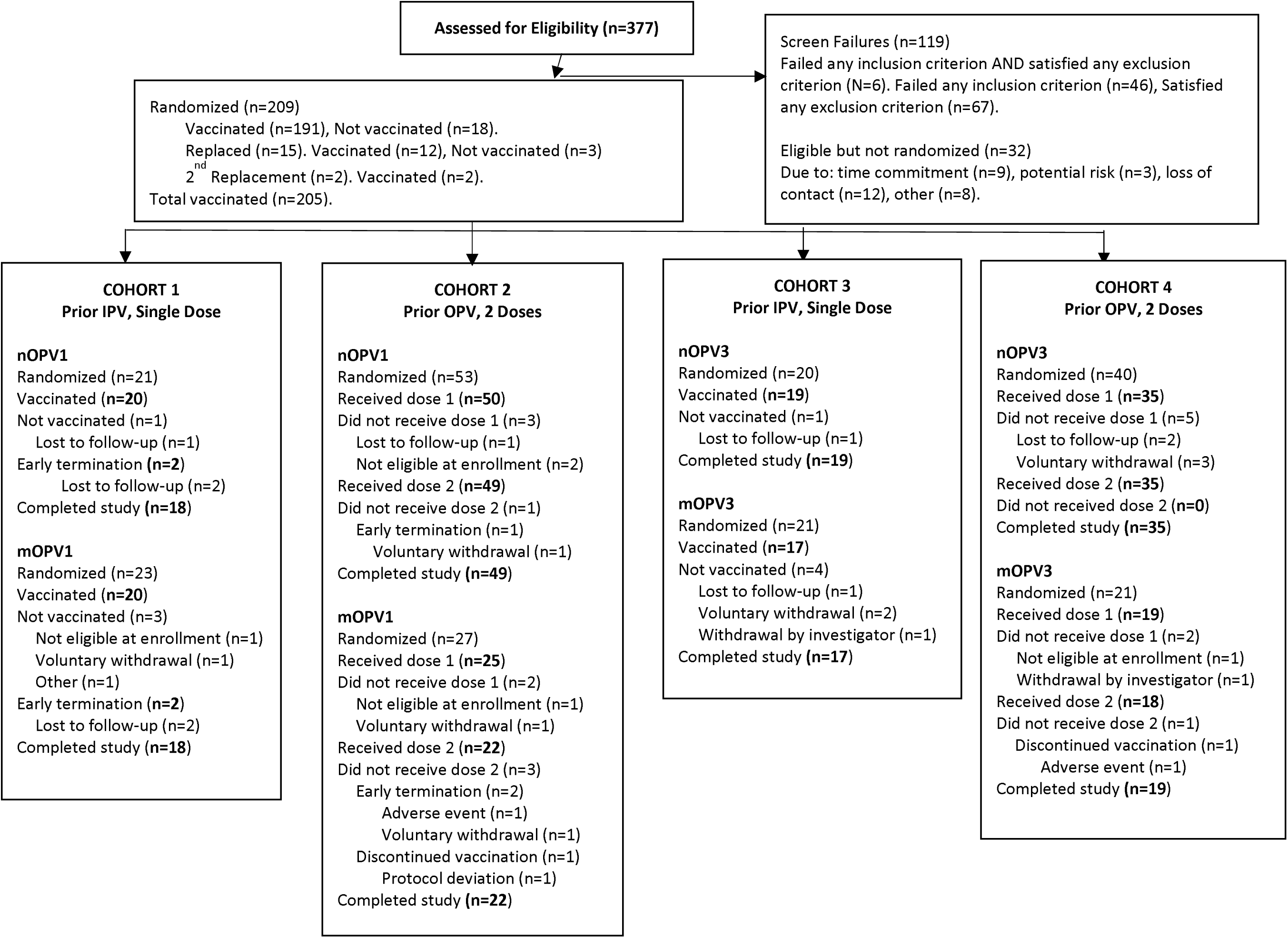
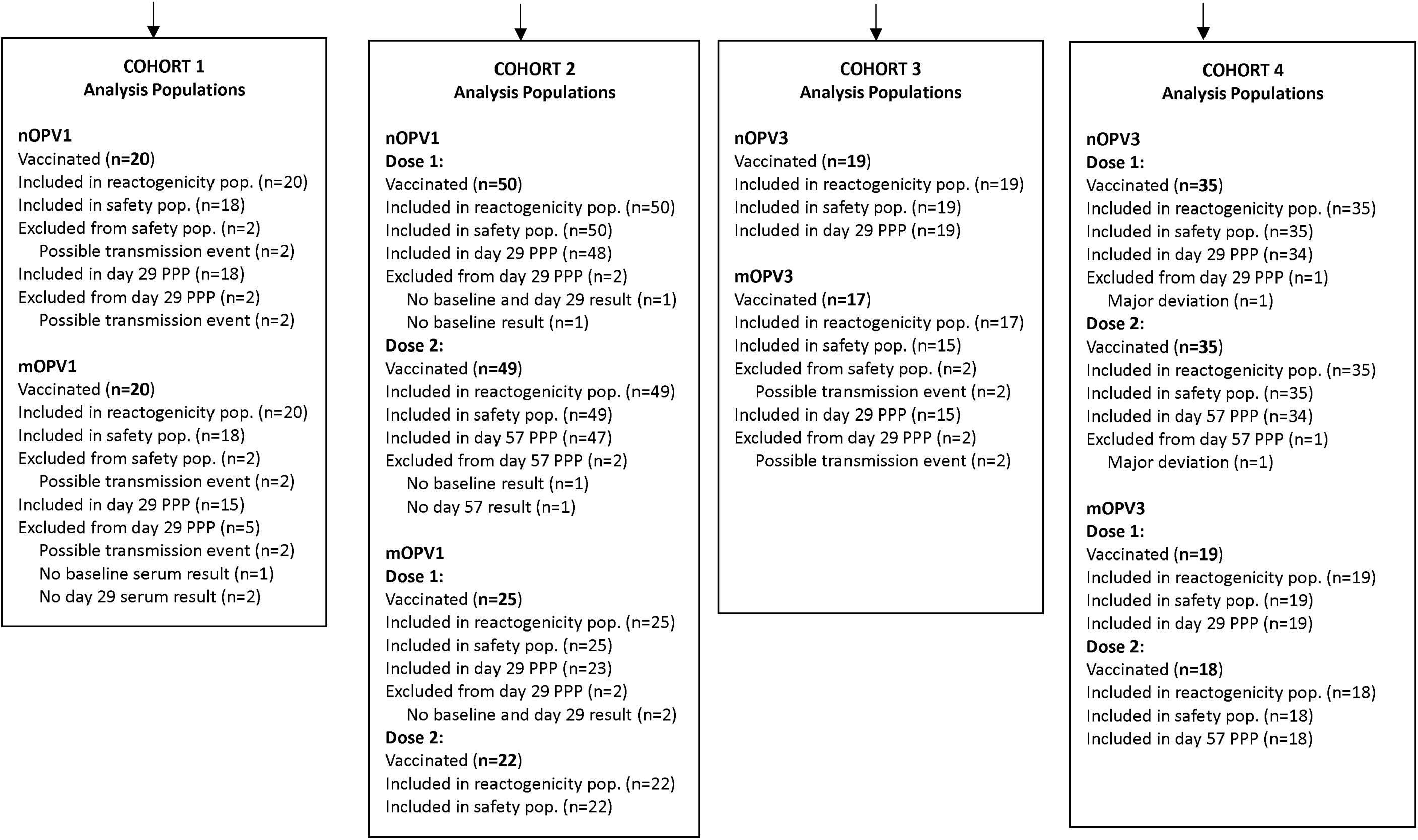
CONSORT diagram

Overall, participants averaged 27·1 years of age, and 55% were female. Among IPV participants (OPV participants), the average age was 20·7 years (30·6 years). Participants were predominantly white, except those enrolled at a single site that enrolled exclusively into cohorts 3 and 4 (due to later study initiation than the other sites), where participants were predominantly African American. Within all cohorts, the nOPV and mOPV groups were balanced in terms of the distribution of sex, ethnicity, race, height, and weight (table 1).

**Table 1.**
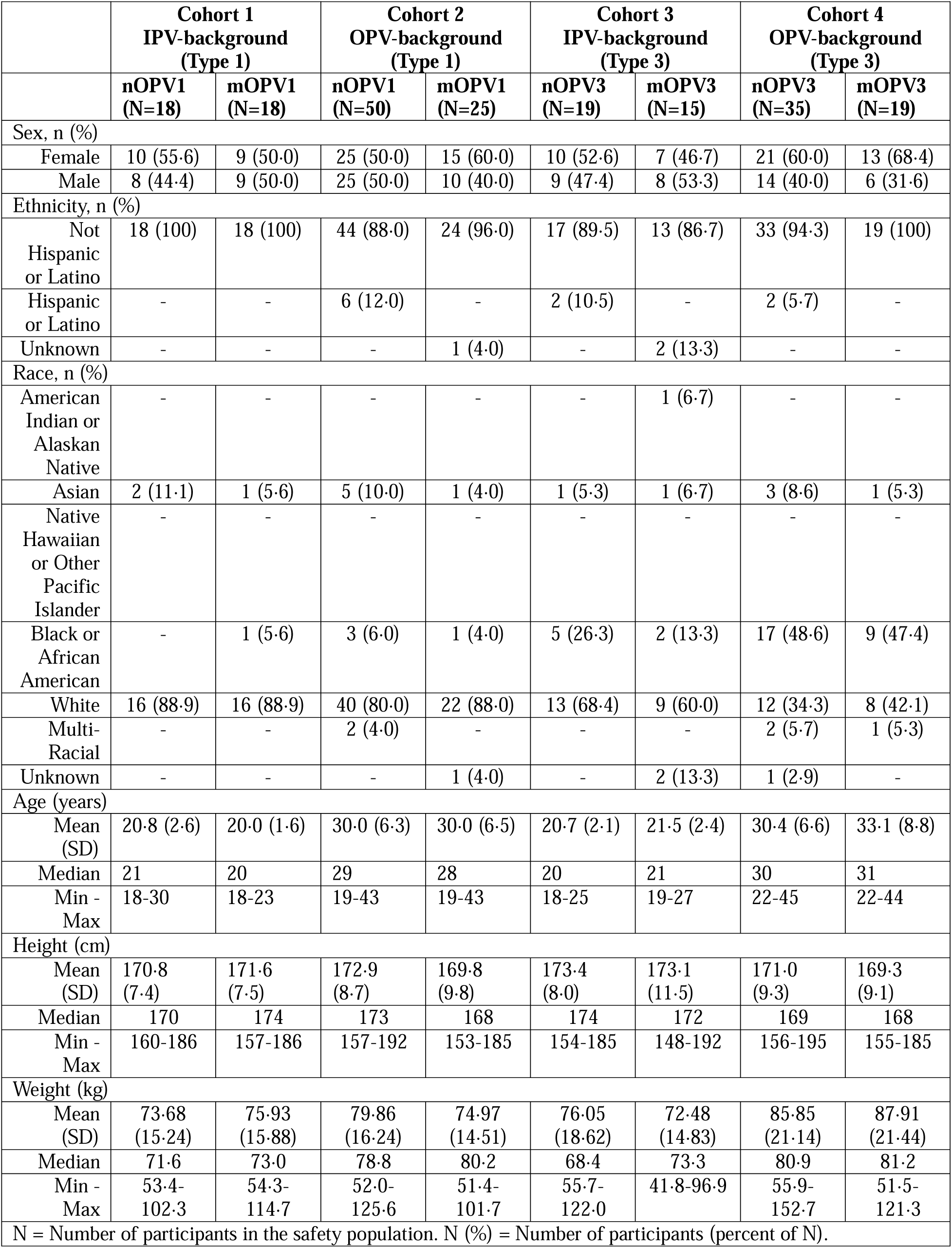
Study group demographics in the Safety Population.

|  | **Cohort 1**<br>**IPV-background**<br>**(Type 1)** |  | **Cohort 2**<br>**OPV-background**<br>**(Type 1)** |  | **Cohort 3**<br>**IPV-background**<br>**(Type 3)** |  | **Cohort 4**<br>**OPV-background**<br>**(Type 3)** |  |
| --- | --- | --- | --- | --- | --- | --- | --- | --- |
|  | **nOPV1**<br>**(N=18)** | **mOPV1**<br>**(N=18)** | **nOPV1**<br>**(N=50)** | **mOPV1**<br>**(N=25)** | **nOPV3**<br>**(N=19)** | **mOPV3**<br>**(N=15)** | **nOPV3**<br>**(N=35)** | **mOPV3**<br>**(N=19)** |
| Sex, n (%) |  |  |  |  |  |  |  |  |
| Female | 10 (55.6) | 9 (50.0) | 25 (50.0) | 15 (60.0) | 10 (52.6) | 7 (46.7) | 21 (60.0) | 13 (68.4) |
| Male | 8 (44.4) | 9 (50.0) | 25 (50.0) | 10 (40.0) | 9 (47.4) | 8 (53.3) | 14 (40.0) | 6 (31.6) |
| Ethnicity, n (%) |  |  |  |  |  |  |  |  |
| Not Hispanic or Latino | 18 (100) | 18 (100) | 44 (88.0) | 24 (96.0) | 17 (89.5) | 13 (86.7) | 33 (94.3) | 19 (100) |
| Hispanic or Latino | - | - | 6 (12.0) | - | 2 (10.5) | - | 2 (5.7) | - |
| Unknown | - | - | - | 1 (4.0) | - | 2 (13.3) | - | - |
| Race, n (%) |  |  |  |  |  |  |  |  |
| American Indian or Alaskan Native | - | - | - | - | - | 1 (6.7) | - | - |
| Asian | 2 (11.1) | 1 (5.6) | 5 (10.0) | 1 (4.0) | 1 (5.3) | 1 (6.7) | 3 (8.6) | 1 (5.3) |
| Native Hawaiian or Other Pacific Islander | - | - | - | - | - | - | - | - |
| Black or African American | - | 1 (5.6) | 3 (6.0) | 1 (4.0) | 5 (26.3) | 2 (13.3) | 17 (48.6) | 9 (47.4) |
| White | 16 (88.9) | 16 (88.9) | 40 (80.0) | 22 (88.0) | 13 (68.4) | 9 (60.0) | 12 (34.3) | 8 (42.1) |
| Multi-Racial | - | - | 2 (4.0) | - | - | - | 2 (5.7) | 1 (5.3) |
| Unknown | - | - | - | 1 (4.0) | - | 2 (13.3) | 1 (2.9) | - |
| Age (years) |  |  |  |  |  |  |  |  |
| Mean (SD) | 20.8 (2.6) | 20.0 (1.6) | 30.0 (6.3) | 30.0 (6.5) | 20.7 (2.1) | 21.5 (2.4) | 30.4 (6.6) | 33.1 (8.8) |
| Median | 21 | 20 | 29 | 28 | 20 | 21 | 30 | 31 |
| Min - Max | 18-30 | 18-23 | 19-43 | 19-43 | 18-25 | 19-27 | 22-45 | 22-44 |
| Height (cm) |  |  |  |  |  |  |  |  |
| Mean (SD) | 170.8 (7.4) | 171.6 (7.5) | 172.9 (8.7) | 169.8 (9.8) | 173.4 (8.0) | 173.1 (11.5) | 171.0 (9.3) | 169.3 (9.1) |
| Median | 170 | 174 | 173 | 168 | 174 | 172 | 169 | 168 |
| Min - Max | 160-186 | 157-186 | 157-192 | 153-185 | 154-185 | 148-192 | 156-195 | 155-185 |
| Weight (kg) |  |  |  |  |  |  |  |  |
| Mean (SD) | 73.68 (15.24) | 75.93 (15.88) | 79.86 (16.24) | 74.97 (14.51) | 76.05 (18.62) | 72.48 (14.83) | 85.85 (21.14) | 87.91 (21.44) |
| Median | 71.6 | 73.0 | 78.8 | 80.2 | 68.4 | 73.3 | 80.9 | 81.2 |
| Min - Max | 53.4-102.3 | 54.3-114.7 | 52.0-125.6 | 51.4-101.7 | 55.7-122.0 | 41.8-96.9 | 55.9-152.7 | 51.5-121.3 |
| N = Number of participants in the safety population. N (%) = Number of participants (percent of N). |  |  |  |  |  |  |  |  |

All 205 participants were included in the reactogenicity population and summaries of solicited events. However, 6 IPV participants, 2 nOPV1 recipients and 2 mOPV1 recipients in cohort 1, and 2 mOPV3 recipients in cohort 3 were removed from the safety and per-protocol populations due to possible transmission of vaccine virus from an alternate study group (identified through NGS of stool samples). Their seven-day post-dose solicited adverse event data are included in table 2, and their unsolicited adverse event data are summarized separately.

**Table 2.** Summary of adverse events post-any dose in the Safety Population.

|  | **Cohorts 1 & 2** |  | **Cohorts 3 & 4** |  |
| --- | --- | --- | --- | --- |
|  | **nOPV1** | **mOPV1** | **nOPV3** | **mOPV3** |
| Number in safety population receiving any dose | 68 | 43 | 54 | 34 |
| Any SAE through the study | 0 (0%) | 0 (0%) | 0 (0%) | 0 (0%) |
| Any AE (solicited or unsolicited) | 50 (74%) | 32 (74%) | 27 (50%) | 20 (59%) |
| Any AE leading to withdrawal from the study | 0 (0%) | 1 (2%) | 0 (0%) | 0 (0%) |
| Severe unsolicited AE within 28 days | 2 (3%) | 1 (2%) | 0 (0%) | 0 (0%) |
| Any unsolicited AE within 28 days | 36 (53%) | 18 (42%) | 9 (17%) | 5 (15%) |
| Any unsolicited AE assessed related to IP within 28 days | 5 (7%) | 2 (5%) | 0 (0%) | 0 (0%) |

Among participants receiving type 1 vaccine (cohorts 1 and 2), no participant reported an SAE and the proportion of participants with any AE (solicited or unsolicited) was similar between the groups: 50 (74%) participants in the nOPV1 groups and 32 (74%) participants in the mOPV1 groups (table 2).

Forty (57%) participants in nOPV1 groups and 31 (69%) participants in mOPV1 groups reported solicited AEs. Most events were mild, with few participants reporting severe events (one each with fatigue and abdominal pain in the nOPV1 groups, and one with severe nausea in the mOPV1 groups). The frequency of the solicited AEs of chills, joint aches/arthralgias, and vomiting were all significantly higher in the mOPV1 group than the nOPV1 groups (p=0.023 for chills, p=0.017 for joint aches/arthralgias, and p=0.022 for vomiting). Chills were reported in 3 (4%) participants in the nOPV1 groups and 8 (18%) participants in the mOPV1 groups, joint aches/arthralgias were reported in 4 (6%) participants in nOPV1 groups and 10 (22%) participants in the mOPV1 groups, and vomiting was reported in none in the nOPV1 groups and 4 (9%) participants in the mOPV1 groups and (table 2). Results were generally similar when limited to within-cohort comparisons (table S6).

Thirty-six (53%) participants in the nOPV1 groups and 18 (42%) participants in the mOPV1 groups reported an unsolicited AE within 28 days post-vaccine administration, most of which were mild. The proportion of participants with any AE assessed by investigators as related was similar in the nOPV1 groups (five [7%] participants) and the mOPV1 groups (two [5%] participants; all were mild except one moderate paresthesia in an mOPV1 recipient). The proportion of participants with any severe AE was similar: two (3%) participants in the nOPV1 groups and one (2%) participant in the mOPV1 groups reported a severe unsolicited AE within 28 days. No participant in the nOPV1 groups and one (2%) participant in the mOPV1 groups reported an AE leading to withdrawal from the study (COVID-19).

Among participants receiving type 3 vaccine, no participant reported an SAE and the proportion of participants with any AE (solicited or unsolicited) was similar between the groups: 27 (50%) participants in the nOPV3 groups and 20 (59%) participants in the mOPV3 groups (table 3).

**Table 3.**
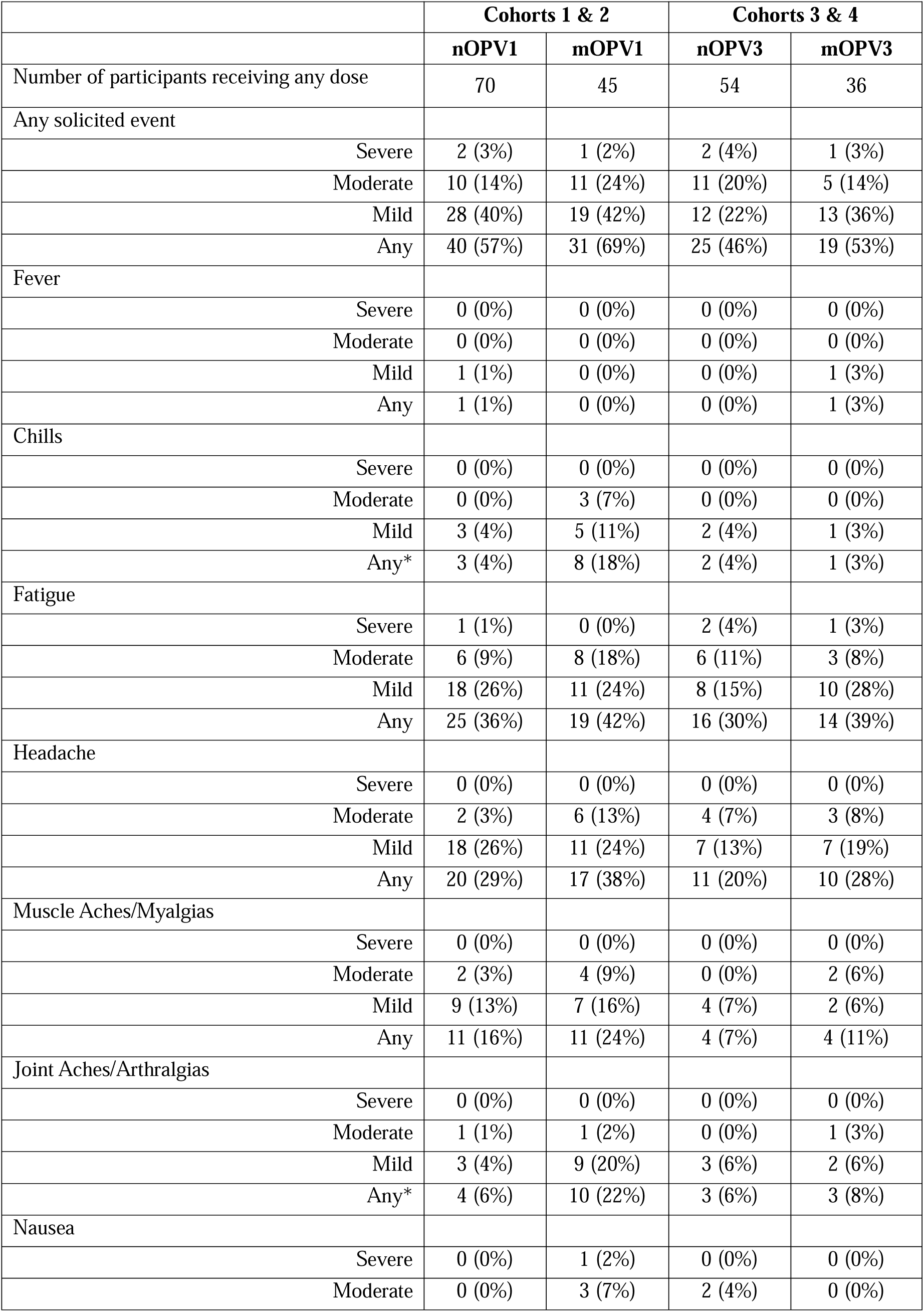
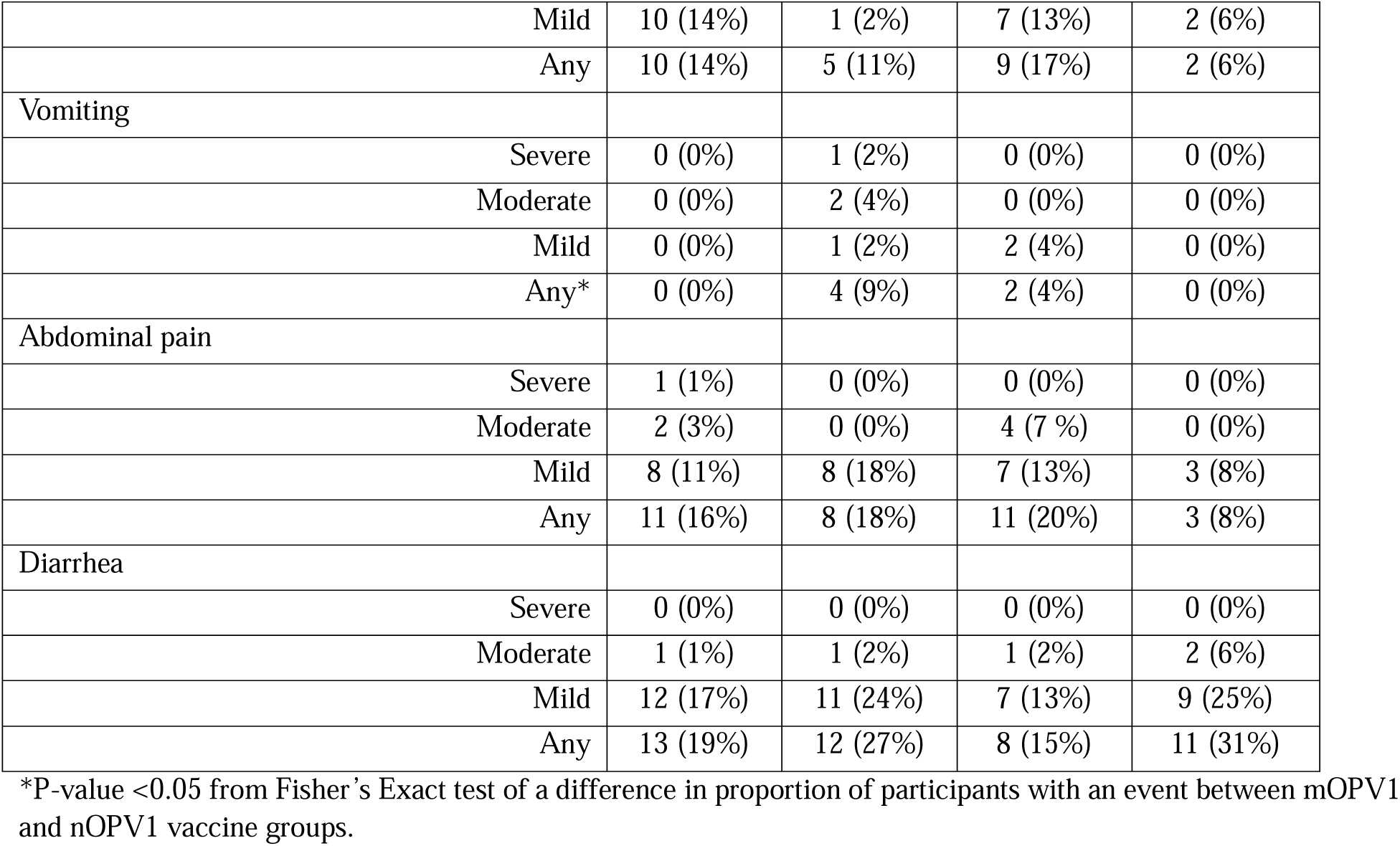
Solicited adverse events after any dose in the Safety Population.

|  | **Cohorts 1 & 2** |  | **Cohorts 3 & 4** |  |
| --- | --- | --- | --- | --- |
|  | **nOPV1** | **mOPV1** | **nOPV3** | **mOPV3** |
| Number of participants receiving any dose | 70 | 45 | 54 | 36 |
| Any solicited event |  |  |  |  |
| Severe | 2 (3%) | 1 (2%) | 2 (4%) | 1 (3%) |
| Moderate | 10 (14%) | 11 (24%) | 11 (20%) | 5 (14%) |
| Mild | 28 (40%) | 19 (42%) | 12 (22%) | 13 (36%) |
| Any | 40 (57%) | 31 (69%) | 25 (46%) | 19 (53%) |
| Fever |  |  |  |  |
| Severe | 0 (0%) | 0 (0%) | 0 (0%) | 0 (0%) |
| Moderate | 0 (0%) | 0 (0%) | 0 (0%) | 0 (0%) |
| Mild | 1 (1%) | 0 (0%) | 0 (0%) | 1 (3%) |
| Any | 1 (1%) | 0 (0%) | 0 (0%) | 1 (3%) |
| Chills |  |  |  |  |
| Severe | 0 (0%) | 0 (0%) | 0 (0%) | 0 (0%) |
| Moderate | 0 (0%) | 3 (7%) | 0 (0%) | 0 (0%) |
| Mild | 3 (4%) | 5 (11%) | 2 (4%) | 1 (3%) |
| Any* | 3 (4%) | 8 (18%) | 2 (4%) | 1 (3%) |
| Fatigue |  |  |  |  |
| Severe | 1 (1%) | 0 (0%) | 2 (4%) | 1 (3%) |
| Moderate | 6 (9%) | 8 (18%) | 6 (11%) | 3 (8%) |
| Mild | 18 (26%) | 11 (24%) | 8 (15%) | 10 (28%) |
| Any | 25 (36%) | 19 (42%) | 16 (30%) | 14 (39%) |
| Headache |  |  |  |  |
| Severe | 0 (0%) | 0 (0%) | 0 (0%) | 0 (0%) |
| Moderate | 2 (3%) | 6 (13%) | 4 (7%) | 3 (8%) |
| Mild | 18 (26%) | 11 (24%) | 7 (13%) | 7 (19%) |
| Any | 20 (29%) | 17 (38%) | 11 (20%) | 10 (28%) |
| Muscle Aches/Myalgias |  |  |  |  |
| Severe | 0 (0%) | 0 (0%) | 0 (0%) | 0 (0%) |
| Moderate | 2 (3%) | 4 (9%) | 0 (0%) | 2 (6%) |
| Mild | 9 (13%) | 7 (16%) | 4 (7%) | 2 (6%) |
| Any | 11 (16%) | 11 (24%) | 4 (7%) | 4 (11%) |
| Joint Aches/Arthralgias |  |  |  |  |
| Severe | 0 (0%) | 0 (0%) | 0 (0%) | 0 (0%) |
| Moderate | 1 (1%) | 1 (2%) | 0 (0%) | 1 (3%) |
| Mild | 3 (4%) | 9 (20%) | 3 (6%) | 2 (6%) |
| Any* | 4 (6%) | 10 (22%) | 3 (6%) | 3 (8%) |
| Nausea |  |  |  |  |
| Severe | 0 (0%) | 1 (2%) | 0 (0%) | 0 (0%) |
| Moderate | 0 (0%) | 3 (7%) | 2 (4%) | 0 (0%) |

|  |  |  |  |  |  |
| --- | --- | --- | --- | --- | --- |
|  | Mild | 10 (14%) | 1 (2%) | 7 (13%) | 2 (6%) |
|  | Any | 10 (14%) | 5 (11%) | 9 (17%) | 2 (6%) |
| Vomiting |  |  |  |  |  |
|  | Severe | 0 (0%) | 1 (2%) | 0 (0%) | 0 (0%) |
|  | Moderate | 0 (0%) | 2 (4%) | 0 (0%) | 0 (0%) |
|  | Mild | 0 (0%) | 1 (2%) | 2 (4%) | 0 (0%) |
|  | Any* | 0 (0%) | 4 (9%) | 2 (4%) | 0 (0%) |
| Abdominal pain |  |  |  |  |  |
|  | Severe | 1 (1%) | 0 (0%) | 0 (0%) | 0 (0%) |
|  | Moderate | 2 (3%) | 0 (0%) | 4 (7 %) | 0 (0%) |
|  | Mild | 8 (11%) | 8 (18%) | 7 (13%) | 3 (8%) |
|  | Any | 11 (16%) | 8 (18%) | 11 (20%) | 3 (8%) |
| Diarrhea |  |  |  |  |  |
|  | Severe | 0 (0%) | 0 (0%) | 0 (0%) | 0 (0%) |
|  | Moderate | 1 (1%) | 1 (2%) | 1 (2%) | 2 (6%) |
|  | Mild | 12 (17%) | 11 (24%) | 7 (13%) | 9 (25%) |
|  | Any | 13 (19%) | 12 (27%) | 8 (15%) | 11 (31%) |

\*P-value <0.05 from Fisher's Exact test of a difference in proportion of participants with an event between mOPV1 and nOPV1 vaccine groups.

None of the differences in the frequency of solicited AEs between the nOPV3 groups and the mOPV3 groups post-any dose were statistically significant. Twenty-five (46%) participants in nOPV3 groups and 19 (53%) participants in mOPV3 groups reported solicited AEs. Most events were mild or moderate, with only a few participants reporting severe events (2 participants in the nOPV3 groups and 1 participant in the mOPV3 groups reported severe fatigue). The most frequently reported moderate events, by participant, were fatigue (6 [11%] in the nOPV3 groups and 3 [8%] in the mOPV3 groups), headache (4 [7%] in the nOPV3 groups and 3 [8%] in the mOPV3 groups), abdominal pain (4 [7%] in the nOPV3 groups and none in the mOPV3 groups), and diarrhea (1 [2%] in the nOPV3 and 2 [6%] in the mOPV3 groups).

Nine (17%) participants in the nOPV3 groups and 5 (15%) participants in the mOPV3 groups reported an unsolicited AE within 28 days post-vaccine administration. No participants receiving nOPV3 or mOPV3 experienced any related AE, severe unsolicited AE within 28 days, or an AE leading to withdrawal.

Among the six participants who were excluded from the post-day-7 safety population due to potential transmission of vaccine virus, one participant in cohort 1 and two in cohort 3 did not report any unsolicited AEs. Only three unsolicited AEs occurred in these participants and were excluded from the safety analyses, including mild muscular weakness, peritonsillar abscess and lymphadenopathy, and moderate back pain and tonsillitis after nOPV1, mild COVID-19 after nOPV1, and mild viral infection after mOPV1.

At baseline across all cohorts, the median neutralizing antibody titres for all three serotypes were slightly lower in the nOPV groups relative to the mOPV groups, except for type 3 in cohort 3 in which baseline titres were similar (table 4, and table S1). In all cohorts, for both nOPV and mOPV, the greatest increases in type-specific neutralizing antibody titres on Day 28 post-dose(s) were homotypic to the vaccine administered, with median (log_2_) values reaching the ULOQ of 10.5 log_2_ by 28 days post dose 1 for type 1 (cohorts 1 and 2) and type 3 (cohorts 3 and 4). High titres were maintained for homotypic virus in all groups in cohorts 2 and 4 at 28 days following the second dose, but there was a slight decline in median titre for type 3 among mOPV3 recipients in cohort 4. While the eligibility screening at Quest demonstrated all participants were seroprotected at baseline, seroprotection rates to poliovirus types 1 and 3 at baseline were less than 100% for all groups when serology testing was conducted at the CDC laboratory, likely attributable to minor assay differences. Strong homotypic responses were observed, with all groups achieving 100% seroprotection 28 days following the first dose.

**Table 4.**
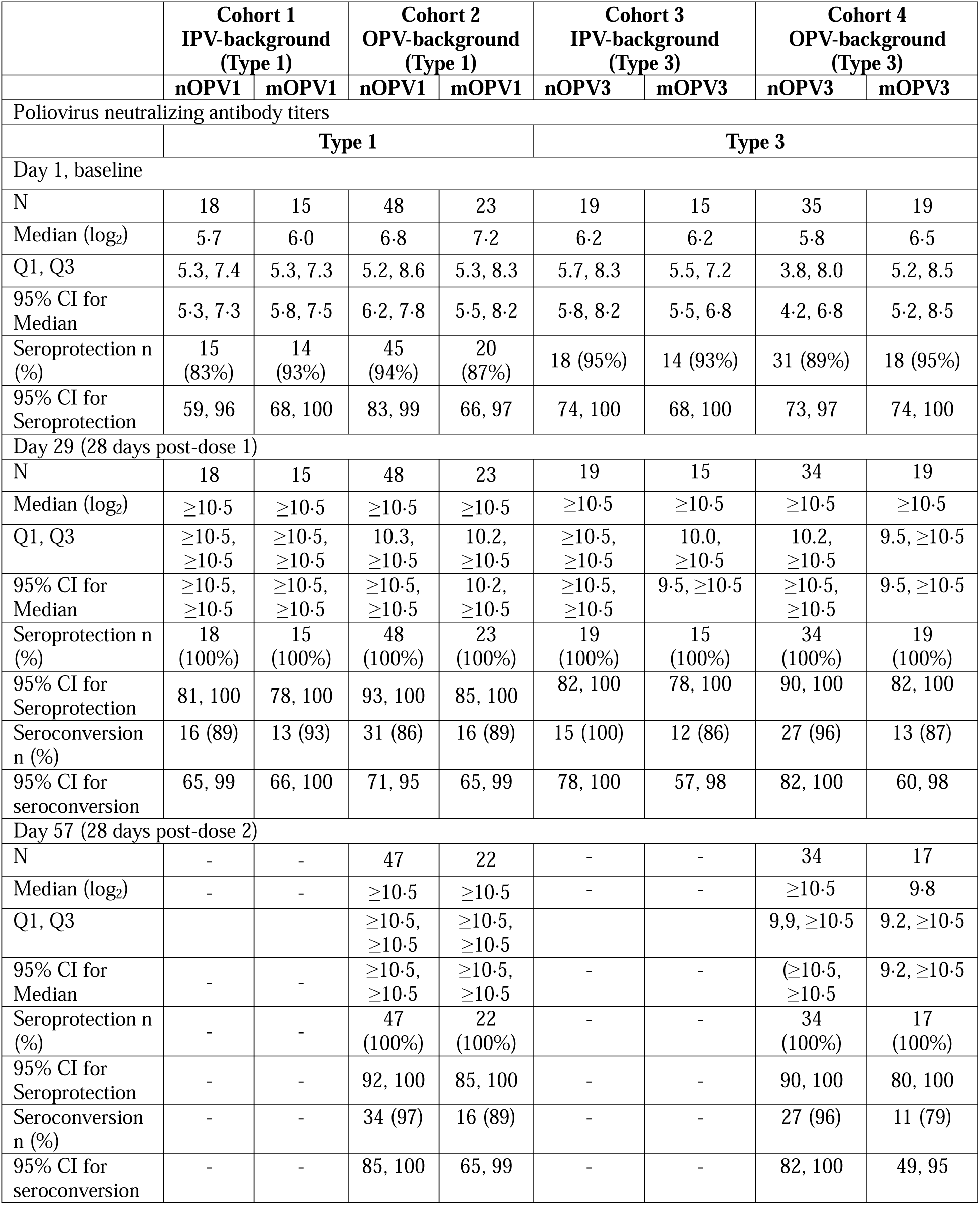
Summaries of homotypic type-specific neutralizing antibody (NAb) titers, seroprotection, and seroconversion rates among the Per-Protocol Population.

Seroconversion rates were high (>85%) in both cohorts 1 and 2 (table 4). Post-dose 2 seroconversion was higher in the nOPV1 group (97%) than the mOPV1 group (89%). In cohorts 3 and 4, seroconversion rates were higher in the nOPV3 groups than in the mOPV3 groups post-dose 1 and post-dose 2 (cohort 4).

Substantial induction of heterotypic immunity against type 2 was observed in both nOPV1 and mOPV1 groups within cohort 2 (OPV participants), with median titres reaching the ULOQ 28 days post-dose 1 (supplementary table 1). A similar heterotypic response was also observed for type 2 among the participants receiving nOPV3 or mOPV3 in cohort 4 (OPV participants). Heterotypic responses to type 2 were observed to a lesser extent in cohort 1 (IPV participants receiving type 1 vaccine) and were not observed in cohort 3 (IPV participants receiving type 3 vaccine), see figure 2. Heterotypic responses between types 1 and 3 were limited among IPV participants and ranged from 7% to 27% following 1 or 2 doses among OPV participants.

**Figure 2:**
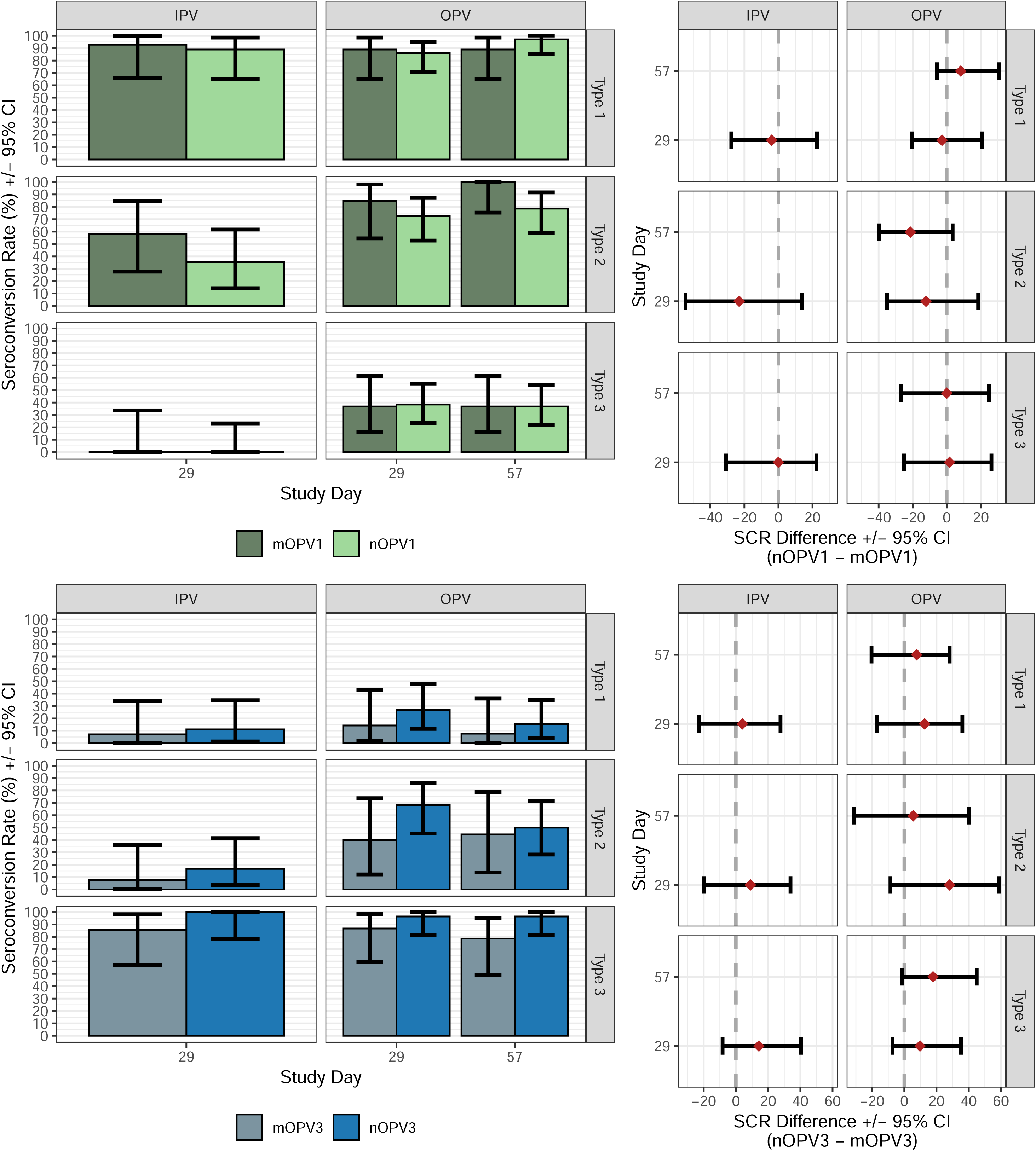
Seroconversion rates (SCR) and differences in seroconversion rates with 95% confidence intervals, by study day and serotype.

The rates of vaccine shedding as measured by PCR in IPV participants in both nOPV and mOPV groups (cohorts 1 and 3) were high (>75%) up to Day 15 post-vaccination, and, except for one mOPV3 recipient, all participants were PCR negative by 7 weeks (figure 3). For OPV participants, rates of viral shedding post-dose 1 were >50% up to Day 15 and <17% by Day 29 (pre-dose 2). Post-dose 2 viral shedding rates in all groups were very low, with the highest rates on Day 36 (7 days post-dose 2): 11% (nOPV1), 10% (mOPV1), 18% (nOPV3), and 6% (mOPV3). Viral infectivity results, as measured by log_10_ CCID_50_/g, demonstrated lower rates, but similar trends (see supplement). PCR results indicate participants in cohorts 1 and 2 shed only poliovirus type 1 and participants in cohorts 3 and 4 shed only poliovirus type 3. As determined by PCR or infectivity, the daily rates and day until cessation of fecal shedding post-dose 1 and post-dose 2 were not meaningfully different between nOPV1 and mOPV1 or between nOPV3 and mOPV3, in any cohort.

**Figure 3.**
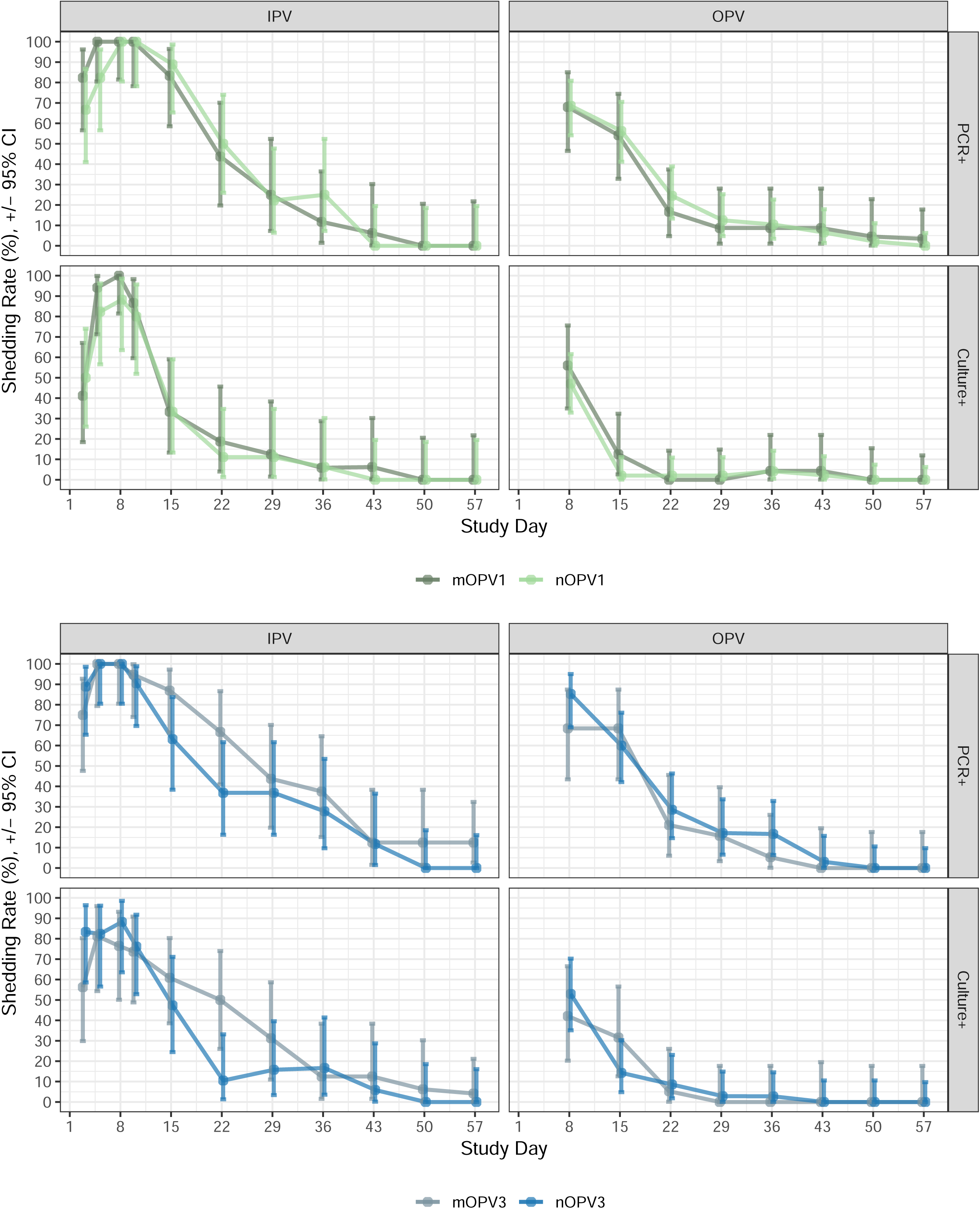
Rate and 95% confidence interval for Type 1 (top) and Type 3 (bottom) vaccine virus shedding positivity as measured by PCR and infectivity.

## Discussion

This first-in-human trial of nOPV1 and nOPV3 demonstrated that both vaccine candidates are safe and well tolerated in healthy adults in the US and can elicit neutralizing antibody responses similar to those elicited by mOPV1 and mOPV3, respectively. In addition, the profile of viral shedding was similar to that of the corresponding mOPV, with shedding in most vaccine recipients limited to the first two weeks post-first vaccination, with subsequent rapid decline to largely undetectable levels by four-to-six weeks post-vaccination. These similar shedding profiles suggest that these chimeric nOPV viruses appear unlikely to be more transmissible.

Of critical importance for polio vaccines is provision of protective immunity as demonstrated by type-specific serum neutralizing antibody levels. Despite demonstration of homotypic seroprotection as an eligibility criterion through commercial laboratory testing, evaluation of immunogenicity outcomes by the CDC using a similar assay measured up to 17% of participants to be seronegative at baseline, depending on the study group. Following a single dose of the nOPV vaccines, all participants demonstrated type-specific protective immunity. The next step for the advancement of these vaccine candidates is demonstration of comparable immunogenicity in geographically relevant target populations of young children and infants.

Detection of vaccine virus in stool provides evidence of successful intestinal infection and viral replication, which is crucial for generation of a robust immune response.(19) Furthermore, reduced vaccine shedding following the second dose in OPV participants indicates successful induction of intestinal immunity, which is critical to halt transmission in outbreak settings.

In addition to robust homotypic serum neutralizing antibody responses, strong heterotypic responses were observed to type 2 poliovirus by both nOPV and mOPV. Such heterotypic responses have been reported in the past, including more frequent heterotypic responses among older age groups, and more frequent type 2 heterotypic responses, compared to types 1 and 3 heterotypic response. (20) More recently, type 1 and 3 responses were observed among IPV-vaccinated children in Lithuania, following administration of type 2 OPV (21), while trivalent OPV-primed children in Pakistan exhibited type 2 responses following mOPV1, and enhanced heterotypic responses following bOPV vaccination. (22) It is possible that heterotypic responses are short-lived as has been observed with other OPV vaccines, however this study does not provide longer-term timepoints necessary for such evaluations.

Limitations of this study include its limited sample size and adult (non-target) population owing to its first-in-human status, as well as the possible transmission events observed.

The value of these candidate vaccines for deployment in the effort to eradicate polio depends not only on demonstration of comparable safety and immunogenicity to their respective mOPVs, but also on demonstration of mucosal immunity for transmission blocking and greater genetic stability with less frequent reversion to neurovirulence. Analysis of next generation sequencing and transgenic mouse neurovirulence testing of fecally shed viruses is ongoing and will be reported elsewhere, including information on the detections resulting from potential transmission events.

The safety and immunogenicity results of this study in healthy adult populations in the US supported the initiation of currently ongoing phase 2 studies in relevant populations of young children and infants, including for nOPV1 alone (ClinicalTrials.gov ID NCT05644184) and combinations of nOPV1, nOPV2, and nOPV3 (ClinicalTrials.gov ID NCT06137664) in Bangladesh, and for nOPV3 in Panama (ClinicalTrials.gov ID NCT05654467), with the aim of achieving licensure and WHO approval for use in outbreak responses.

## Contributors

LDM, ASB, AF, JOK, and CG participated in the conception and design of the study and LDM, AF, JOK and CG contributed to the writing of the protocol. ACS, ERC, JWC, PFW, and MAI were site investigators. AV, BAM, and YZ were responsible for all viral analyses and assessments of the neutralizing antibodies. JLK-A provided interpretation of virological results.

ET supplied the vaccines. All authors contributed to the interpretation of the results. LM and AF accessed and verified the data and led the writing of the initial drafts of the manuscript on which all authors commented, and all authors had full access to all the data and had final responsibility for the decision to submit for publication.

## Data sharing

Data for this study will be made available to others in the scientific community upon request. Standard criteria for making data available for valid research projects will be used, following application by suitably qualified researchers. For data access, please contact the corresponding author.

## Acknowledgements

The authors are grateful to study participants, study site staff members, and other site investigators (including Ian Carroll and Luther Bartelt at UNC-Chapel Hill) for their contributions to the study. We thank Carrie Trujillo, Rahsan Erdem, Renee Holt, Mike Raine, and Shadia Lopez for their efforts in implementing the study and Abdi Naficy for contributions to design of the study. We thank the laboratory staff that performed the serology testing (Basit Jafri, Kathryn Jones, William Hendley, and Sandra Valdez) and shedding testing (Talha Abid, Ray Campagnoli, Elizabeth Coffee, Larin McDuffie, Esther Morantz, Lindsey Oteyza, Ling Wei, and Amanda Williams) at the Polio and Picornavirus Branch and the Specimen Triage and Tracking Team at the CDC. We are grateful to Drs. Andrew Macadam and Raul Andino and their groups at MHRA and UCSF for developing the nOPV vaccine candidates and conducting critical pre-clinical research which enabled this trial.

## References

1. GPEI. History of Polio [Available from: https://polioeradication.org/polio-today/history-of-polio/.

2. WHO. Polio Eradication Strategy 2022–2026: Delivering on a promise. 2021.

3. McCarthy KA, Chabot-Couture G, Famulare M, Lyons HM, Mercer LD. The risk of type 2 oral polio vaccine use in post-cessation outbreak response. BMC medicine. 2017;15(1):175.

4. Gray EJ, Cooper LV, Bandyopadhyay AS, Blake IM, Grassly NC. The Origins and Risk Factors for Serotype-2 Vaccine-Derived Poliovirus Emergences in Africa During 2016-2019. The Journal of infectious diseases. 2023;228(1):80–8.

5. Van Damme P, De Coster I, Bandyopadhyay AS, Revets H, Withanage K, De Smedt P, et al. The safety and immunogenicity of two novel live attenuated monovalent (serotype 2) oral poliovirus vaccines in healthy adults: a double-blind, single-centre phase 1 study. Lancet. 2019;394(10193):148–58.

6. De Coster I, Leroux-Roels I, Bandyopadhyay AS, Gast C, Withanage K, Steenackers K, et al. Safety and immunogenicity of two novel type 2 oral poliovirus vaccine candidates compared with a monovalent type 2 oral poliovirus vaccine in healthy adults: two clinical trials. Lancet. 2021;397(10268):39–50.

7. Saez-Llorens X, Bandyopadhyay AS, Gast C, Leon T, DeAntonio R, Jimeno J, et al. Safety and immunogenicity of two novel type 2 oral poliovirus vaccine candidates compared with a monovalent type 2 oral poliovirus vaccine in children and infants: two clinical trials. Lancet. 2021;397(10268):27–38.

8. WHO. First ever vaccine listed under WHO emergency use [Available from: https://www.who.int/news/item/13-11-2020-first-ever-vaccine-listed-under-who-emergency-use.

9. Ochoge M, Futa AC, Umesi A, Affleck L, Kotei L, Daffeh B, et al. Safety of the novel oral poliovirus vaccine type 2 (nOPV2) in infants and young children aged 1 to <5 years and lot-to-lot consistency of the immune response to nOPV2 in infants in The Gambia: a phase 3, double-blind, randomised controlled trial. Lancet. 2024;403(10432):1164–75.

10. Zaman K, Bandyopadhyay AS, Hoque M, Gast C, Yunus M, Jamil KM, et al. Evaluation of the safety, immunogenicity, and faecal shedding of novel oral polio vaccine type 2 in healthy newborn infants in Bangladesh: a randomised, controlled, phase 2 clinical trial. Lancet. 2022.

11. Bandyopadhyay AS, Cooper LV, Zipursky S. One billion doses and WHO prequalification of nOPV2: Implications for the global polio situation and beyond. PLOS Glob Public Health. 2024;4(2):e0002920.

12. Voorman A, Lyons H, Shuaib F, Adamu US, Korir C, Erbeto T, et al. Impact of Supplementary Immunization Activities using Novel Oral Polio Vaccine Type 2 during a Large outbreak of Circulating Vaccine-Derived Poliovirus in Nigeria. The Journal of infectious diseases. 2024;229(3):805–12.

13. Martin J, Burns CC, Jorba J, Shulman LM, Macadam A, Klapsa D, et al. Genetic Characterization of Novel Oral Polio Vaccine Type 2 Viruses During Initial Use Phase Under Emergency Use Listing - Worldwide, March-October 2021. MMWR Morbidity and mortality weekly report. 2022;71(24):786–90.

14. Peak CM, Lyons H, Voorman A, Gray EJ, Cooper LV, Blake IM, et al. Monitoring the Risk of Type-2 Circulating Vaccine-Derived Poliovirus Emergence During Roll-Out of Type-2 Novel Oral Polio Vaccine. Vaccines. 2024;12(12).

15. Yeh MT, Smith M, Carlyle S, Konopka-Anstadt JL, Burns CC, Konz J, et al. Genetic stabilization of attenuated oral vaccines against poliovirus types 1 and 3. Nature. 2023;619(7968):135-42.

16. Yeh MT, Bujaki E, Dolan PT, Smith M, Wahid R, Konz J, et al. Engineering the Live-Attenuated Polio Vaccine to Prevent Reversion to Virulence. Cell host & microbe. 2020;27(5):736–51 e8.

17. FDA. 2007 [Available from: chrome-extension://efaidnbdmnnnibpcajpcglclefindmkaj/https://www.fda.gov/media/73679/download.

18. Weldon CW, Oberste MS, Pallansch MA. Standardization Methods for Detecton of Poliovirus Antibodies. In: Media sSB, editor. Poliovirus: Methods and Protocols2016. p. 145–76.

19. Hird TR, Grassly NC. Systematic review of mucosal immunity induced by oral and inactivated poliovirus vaccines against virus shedding following oral poliovirus challenge. PLoS pathogens. 2012;8(4):e1002599.

20. Ashkenazi A, Melnick JL. Heterotypic antibody response after feeding of monovalent attenuated live poliovaccine. The New England journal of medicine. 1962;267:1228–30.

21. Bandyopadhyay AS, Gast C, Brickley EB, Ruttimann R, Clemens R, Oberste MS, et al. A Randomized Phase 4 Study of Immunogenicity and Safety After Monovalent Oral Type 2 Sabin Poliovirus Vaccine Challenge in Children Vaccinated with Inactivated Poliovirus Vaccine in Lithuania. The Journal of infectious diseases. 2021;223(1):119–27.

22. Mir F, Quadri F, Mach O, Ahmed I, Bhatti Z, Khan A, et al. Monovalent type-1 oral poliovirus vaccine given at short intervals in Pakistan: a randomised controlled, four-arm, open-label, non-inferiority trial. The Lancet Infectious diseases. 2015;15(8):889–97.

