## Supplementary Material for "First-in-human, phase 1, randomized, observer-blind, controlled trial to assess the safety and immunogenicity of novel live attenuated type 1 and type 3 oral poliomyelitis vaccines in healthy adults"

### Description of potential poliovirus transmission observed during the study

Due to the different study products in use and unique, intensive stool collection and testing performed in this study, an opportunity to observe potential OPV transmission events was created. This section summarizes the observations made in this study.

Potential transmission events were identified in six participants with IPV-only vaccination history (2 nOPV1 recipients and 2 mOPV1 recipients in cohort 1, and 2 mOPV3 recipients in cohort 3). These participants were excluded from the Safety and Per-protocol Populations (but not the Reactogenicity Population) due to the genomic detection (PCR and/or NGS testing) in at least one shed stool sample of the alternate vaccine used within the respective cohort. In one of the six cases, a mixture of both mOPV1 and nOPV1 was detected.

Following an investigation by the study team, including additional analysis of the impacted samples, two main hypotheses for the root cause were deemed most probable: 1) limited, inadvertent cross-contamination of stool samples during handling, processing, or testing of the impacted stool specimens, or 2) possible transmission events between study participants who were in close contact (e.g., roommates, romantic partners, teammates, close acquaintances, etc.) and had received opposing vaccines.

For five of the six excluded participants, only a single stool sample or single stool aliquot demonstrated the unanticipated alternate vaccine profile, suggesting inadvertent cross-contamination during stool handling may have occurred. Notably, the extremely sensitive molecular techniques employed in this study could detect very low levels of cross-sample contamination.

For the remaining one of six excluded participants, a nOPV1 recipient, multiple stool samples demonstrated the presence of mOPV1 or a mixture of both mOPV1 and nOPV1, suggesting sample contamination alone is an unlikely explanation. During investigation, this study participant reported sustained close contact with another study participant who was determined to have received the alternate study vaccine. As such, a transmission event was deemed to be the most probable root cause.

For all six participants, it remains difficult to fully rule out transmission as a possibility. Therefore, all six situations were conservatively treated as “potential transmission events” and all six participants were excluded from the safety and per-protocol populations.

### **S. Table 1:** Summaries of heterotypic type-specific neutralizing antibody (NAb) titres measured at baseline and 28 days post each dose among the Per-Protocol Population.

|  | **Cohort 1**  **IPV-background (Type 1)** | | **Cohort 2**  **OPV-background**  **(Type 1)** | | **Cohort 3**  **IPV-background**  **(Type 3)** | | **Cohort 4**  **OPV-background**  **(Type 3)** | |
| --- | --- | --- | --- | --- | --- | --- | --- | --- |
|  | **nOPV1** | **mOPV1** | **nOPV1** | **mOPV1** | **nOPV3** | **mOPV3** | **nOPV3** | **mOPV3** |
| **Poliovirus neutralizing antibody titres** | | | | | | | | |
|  | **Type 3** | | | | **Type 1** | | | |
| Day 1, baseline | | | | | | | | |
| N | 18 | 15 | 48 | 23 | 19 | 15 | 35 | 19 |
| Median (log_2_) | 8·0 | 8·2 | 6·0 | 6·5 | 4·8 | 5·2 | 6·5 | 6·8 |
| 95% CI for Median | 4·8, 8·5 | 5·5, 9·5 | 5·0, 7·5 | 3·5, 7·5 | 4·2, 7·2 | 3·8, 6·8 | 5·5, 8·5 | 5·2, 8·2 |
| Seroprotection n (%) | 14 (78%) | 13 (87%) | 44 (92%) | 20 (87%) | 18 (95%) | 14 (93%) | 33 (94%) | 16 (84%) |
| 95% CI for Seroprotection | 52, 94 | 60, 98 | 80, 98 | 66, 97 | 74, 100 | 68, 100 | 81, 99 | 60, 97 |
| Day 29 (28 days post-dose 1) | | | | | | | | |
| N | 18 | 15 | 48 | 23 | 19 | 15 | 34 | 19 |
| Median (log_2_) | 7·3 | 7·5 | 8·5 | 8·2 | 6·2 | 5·5 | 7·0 | 7·2 |
| 95% CI for Median | 5·2, 8·8 | 5·2, 9·8 | 7·5, 9·2 | 7·5, 9·5 | 4·5, 7·8 | 3·8, 6·8 | 6·2, 9·2 | 5·8, 8·5 |
| Seroprotection n (%) | 14 (78%) | 13 (87%) | 44 (92%) | 22 (96%) | 17 (90%) | 15 (100%) | 32 (94%) | 17 (90%) |
| 95% CI for Seroprotection | 52, 94 | 60, 98 | 80, 98 | 78, 100 | 67, 99 | 78, 100 | 80, 99 | 67, 99 |
| Day 57 (28 days post-dose 2) | | | | | | | | |
| N | - | - | 47 | 22 | - | - | 34 | 17 |
| Median (log_2_) | - | - | 8·2 | 7·8 | - | - | 7·7 | 6·8 |
| 95% CI for Median | - | - | 6·8, 8·5 | 6·2, 8·8 | - | - | 5·8, 9·2 | 4·8, 8·2 |
| Seroprotection n (%) | - | - | 45 (96%) | 21 (96%) | - | - | 31 (91%) | 15 (88%) |
| 95% CI for Seroprotection | - | - | 85, 99 | 77, 100 | - | - | 76, 98 | 64, 99 |
| **Type 2** | | | | | | | | |
| Day 1, baseline | | | | | | | | |
| N | 18 | 15 | 48 | 23 | 19 | 15 | 35 | 19 |
| Median (log_2_) | 7·2 | 7·5 | 7·5 | 8·2 | 5·8 | 6·2 | 7·8 | 8·5 |
| 95% CI for Median | 5·0, 7·7 | 5·5, 8·2 | 6·5, 9·0 | 6·2, 9·8 | 4·5, 7·5 | 3·8, 8·5 | 6·5, 8·8 | 6·5, 9·8 |
| Seroprotection n (%) | 15 (83%) | 14 (93%) | 48 (100%) | 23 (100%) | 19 (100%) | 15 (100%) | 32 (91%) | 19 (100%) |
| 95% CI for Seroprotection | 59, 96 | 68, 100 | 93, 100 | 85, 100 | 82, 100 | 78, 100 | 77, 98 | 82, 100 |
| Day 29 (28 days post-dose 1) | | | | | | | | |
| N | 18 | 15 | 48 | 23 | 19 | 15 | 34 | 19 |
| Median (log_2_) | 7·8 | 8·8 | ≥10·5 | ≥10·5 | 5·8 | 6·2 | 10·2 | 10·2 |
| 95% CI for Median | 6.8, 9.3 | 7.5, ≥10.5) | ≥10.5, ≥10.5 | ≥10.5, ≥10.5 | 4.8, 8.5 | 3.5, 8.2 | 9.7, 10.2 | 8.2, ≥10.5 |
| Seroprotection n (%) | 17 (94%) | 15 (100%) | 48 (100%) | 23 (100%) | 19 (100%) | 13 (87%) | 33 (97%) | 18 (95%) |
| 95% CI for Seroprotection | 73, 100 | 78, 100 | 93, 100 | 85, 100 | 82, 100 | 60, 98 | 85, 100 | 74, 100 |
| Day 57 (28 days post-dose 2) | | | | | | | | |
| N | - | - | 47 | 22 | - | - | 34 | 17 |
| Median (log_2_) | - | - | ≥10.5 | ≥10.5 | - | - | 9.7 | 9.5 |
| 95% CI for Median | - | - | 10.2, ≥10.5 | 10.2, ≥10.5 | - | - | 8.5, 10.2 | 8.8, 10.2 |
| Seroprotection n (%) | - | - | 47 (100%) | 22 (100%) | - | - | 33 (97%) | 17 (100%) |
| 95% CI for Seroprotection | - | - | 92, 100 | 85, 100 | - | - | 85, 100 | 80, 100 |

### **S. Table 2:** Heterotypic type-specific seroconversion rates measured by neutralizing antibody titres from baseline, in Per Protocol Population participants for which it is possible to observe seroconversion (log_2_ titre ≤8·5 at baseline).

|  | **Cohort 1**  **IPV-background (Type 1)** | | **Cohort 2**  **OPV-background**  **(Type 1)** | | **Cohort 3**  **IPV-background**  **(Type 3)** | | **Cohort 4**  **OPV-background**  **(Type 3)** | |
| --- | --- | --- | --- | --- | --- | --- | --- | --- |
|  | **nOPV1** | **mOPV1** | **nOPV1** | **mOPV1** | **nOPV3** | **mOPV3** | **nOPV3** | **mOPV3** |
| Seroconversion | | | | | | | | |
|  | **Type 3** | | | | **Type 1** | | | |
| Day 29, post-dose 1 | | | | | | | | |
| N | 14 | 9 | 39 | 19 | 18 | 14 | 26 | 14 |
| n (%) | 0 (0) | 0 (0) | 15 (39) | 7 (37) | 2 (11) | 1 (7) | 7 (27) | 2 (14) |
| 95% CI | (0, 23) | (0, 34) | (23, 55) | (16, 62) | (1, 35) | (0, 34) | (12, 48) | (2, 43) |
| N | - | - | 38 | 19 | - | - | 26 | 14 |
| n (%) | - | - | 14 (37) | 7 (37) | - | - | 4 (15) | 1 (7) |
| 95% CI | - | - | (22, 54) | (16, 62) | - | - | (4, 35) | (0, 34) |
| ***Type 2*** | | | | | | | | |
| Day 29, post-dose 1 | | | | | | | | |
| N | 17 | 12 | 29 | 13 | 18 | 13 | 22 | 10 |
| n (%) | 6 (35) | 7 (58) | 21 (72) | 11 (85) | 3 (17) | 1 (8) | 15 (68) | 4 (40) |
| 95% CI | (14, 62) | (28, 85) | (53, 87) | (55, 98) | (4, 41) | (0, 36) | (45, 86) | (12, 74) |
| Day 57, post-dose 2 | | | | | | | | |
| N | - | - | 28 | 13 | - | - | 22 | 10 |
| n (%) | - | - | 22 (79) | 13 (100) | - | - | 11 (50) | 4 (40) |
| 95% CI | - | - | (59, 92) | (75, 100) | - | - | (28, 72) | (12, 74) |

**S. Figure 1:** Geometric mean titres (GMTs) with 95% CIs by study day and serotype.
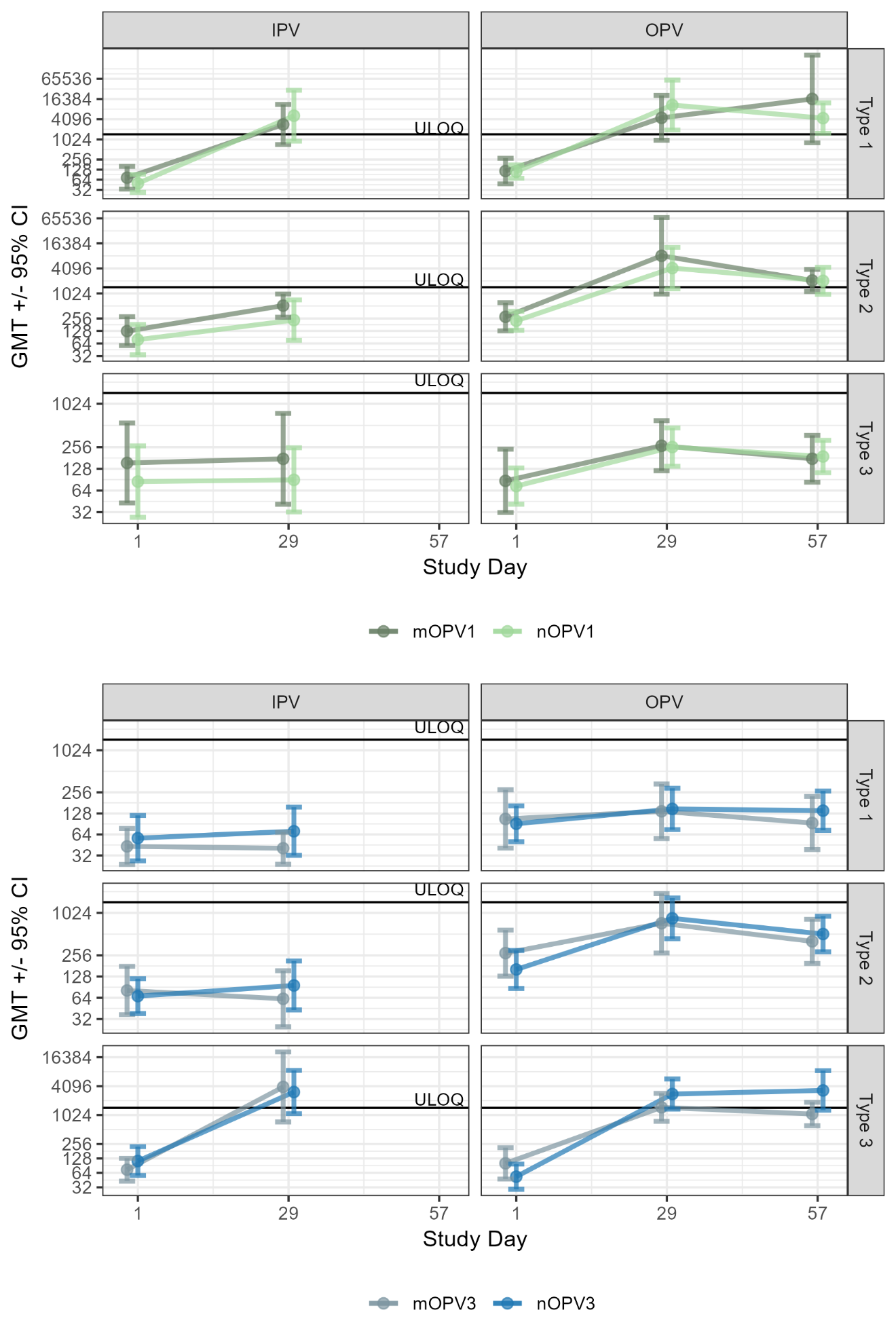

### **S. Table 3:** Time (days) to cessation of fecal shedding in IPV participants (cohorts 1 and 3) in the Safety Population.

| Method/ Cohort | Quartile | nOPV | | mOPV | |  |
| --- | --- | --- | --- | --- | --- | --- |
|  |  | Estimate | 95% CI | Estimate | 95% CI ^a^ | P-value ^b^ |
| **PCR** | | | | | | |
| Cohort 1 (Type 1) | 25% | 20 | 15 - 22 | 20 | 14 - 20 |  |
|  | 50% (Median) | 22 | 15 - 24 | 20 | 20 - 29 | 0·975 |
|  | 75% | 29 | 22 - 42 | 29 | 20 - 43 |  |
| Cohort 3 (Type 3) | 25% | 20 | 10 - 20 | 22 | 13 - 29 |  |
|  | 50% (Median) | 20 | 15 - 37 | 29 | 22 - 42 | 0·086 |
|  | 75% | 37 | 20 - 50 | 42 | 29 - 79 |  |
| **Culture (> 2·75 log_10_ CCID_50_/g)** | | | | | | |
| Cohort 1 (Type 1) | 25% | 10 | 8 - 15 | 13 | 9 - 13 |  |
|  | 50% (Median) | 15 | 10 - 15 | 13 | 13 - 16 | 0·683 |
|  | 75% | 16 | 15 - 36 | 16 | 13 - 43 |  |
| Cohort 3 (Type 3) | 25% | 14 | 2 - 15 | 13 | 2 - 22 |  |
|  | 50% (Median) | 15·5 | 14 - 20 | 25 | 13 - 35 | 0·220 |
|  | 75% | 20 | 15 - 42 | 35 | 22 - 43 |  |
| **Culture (≥ 4·0 log_10_ CCID_50_/g)** | | | | | | |
| Cohort 1 (Type 1) | 25% | 2 | NC | 9 | 2 - 11 |  |
|  | 50% (Median) | 9 | 2 - 11 | 11 | 9 - 11 | 0·158 |
|  | 75% | 11 | 9 - 24 | 11 | NC |  |
| Cohort 3 (Type 3) | 25% | 5 | 2 - 8 | 6 | 2 - 9 |  |
|  | 50% (Median) | 13 | 5 - 13 | 9 | 2 - 14 | 0·941 |
|  | 75% | 13 | 13 - 16 | 14 | 9 - 28 |  |
| NC = Not calculated ^a^ 95% confidence interval calculated utilizing interval-censoring methodology. ^b^ P-value calculated from the generalized log-rank test. | | | | | | |

### **S. Table 4:** Shedding Index Endpoint (SIE), by cohort, dose, and vaccine received in the Safety Population.

|  | | SIE | | | Difference (nOPV – mOPV) | | |
| --- | --- | --- | --- | --- | --- | --- | --- |
| Cohort (Prior Vaccination) | Vaccination Group | n | Median | 95% CI | Estimate | 95% CI | P-Value |
| **Post-Dose 1** | | | | | | | |
| Cohort 1 (IPV) | nOPV1 | 16 | 1·88 | 1·477, 2·344 | -0·08 | -0·828, 0·484 | 0·614 |
|  | mOPV1 | 13 | 1·96 | 1·734, 2·477 |  |  |  |
| Cohort 2 (OPV) | nOPV1 | 45 | 1·38 | 0·688, 1·484 | -0·09 | -0·992, 0·238 | 0·422 |
|  | mOPV1 | 17 | 1·46 | 0·719, 1·695 |  |  |  |
| Cohort 3 (IPV) | nOPV3 | 14 | 2·20 | 1·844, 3·016 | -0·21 | -1·484, 0·891 | 0·443 |
|  | mOPV3 | 11 | 2·41 | 1·766, 3·578 |  |  |  |
| Cohort 4 (OPV) | nOPV3 | 24 | 1·38 | 0·688, 1·539 | -0·07 | -0·930, 0·695 | 0·878 |
|  | mOPV3 | 16 | 1·44 | 0·688, 2·055 |  |  |  |
| **Post-Dose 2** | | | | | | | |
| Cohort 2 (OPV) | nOPV1 | 34 | 0·00 | 0·000, 0·000 | 0·00 | 0·000, 0·000 | 0·960 |
|  | mOPV1 | 19 | 0·00 | 0·000, 0·000 |  |  |  |
| Cohort 4 (OPV) | nOPV3 | 26 | 0·00 | 0·000, 0·000 | 0·00 | 0·000, 0·000 | 0·245 |
|  | mOPV3 | 13 | 0·00 | 0·000, 0·000 |  |  |  |
| N = Number of subjects vaccinated and with samples collected (delivered to the site) at all 4 time points.  95% CI = Bootstrap-based 95% confidence interval for the median and difference in medians (10,000 samples). P-Value = Wilcoxon 2-sample test of a difference between groups. SIE is calculated as the arithmetic mean of log10 CCID50/g across the appropriate nominal days or within respective sampling windows.  Log10 CCID50 per gram ≤LLOQ contributed a value equal to the LLOQ. | | | | | | | |

### **S. Table 5:** Viral shedding Area Under the Curve (AUC), by cohort, dose, and vaccine received in the Safety Population.

|  | | AUC | | | Difference (nOPV – mOPV) | | |
| --- | --- | --- | --- | --- | --- | --- | --- |
| Cohort (Prior Vaccination) | Vaccination Group | n | Median | 95% CI | Estimate | 95% CI | P-Value |
| **Post-Dose 1 Days 7 to 28** | | | | | | | |
| Cohort 2 (OPV) | nOPV1 | 46 | 27·30 | 9·625, 31·125 | -2·05 | -22·945, 17·359 | 0·689 |
|  | mOPV1 | 18 | 29·35 | 9·156, 35·625 |  |  |  |
| Cohort 4 (OPV) | nOPV3 | 26 | 27·09 | 9·625, 30·766 | -5·28 | -24·125, 11·000 | 0·566 |
|  | mOPV3 | 17 | 32·38 | 17·875, 38·500 |  |  |  |
| **Post-Dose 1 Days 2 to 28** | | | | | | | |
| Cohort 1 (IPV) | nOPV1 | 15 | 57·84 | 40·188, 66·625 | -9·03 | -33·688, 6·313 | 0·229 |
|  | mOPV1 | 15 | 66·88 | 52·797, 78·344 |  |  |  |
| Cohort 3 (IPV) | nOPV3 | 17 | 60·80 | 49·172, 80·438 | -10·66 | -32·406, 14·941 | 0·319 |
|  | mOPV3 | 12 | 71·46 | 56·711, 87·945 |  |  |  |
| **Post-Dose 1 Days 2 to 56** | | | | | | | |
| Cohort 1 (IPV) | nOPV1 | 14 | 58·43 | 42·086, 66·625 | -6·20 | -44·797, 14·398 | 0·482 |
|  | mOPV1 | 13 | 64·63 | 51·531, 96·359 |  |  |  |
| Cohort 3 (IPV) | nOPV3 | 16 | 72·37 | 49·359, 100·734 | -17·77 | -61·602, 30·969 | 0·501 |
|  | mOPV3 | 12 | 90·14 | 56·711, 122·156 |  |  |  |
| **Post-Dose-2 Days 28 to 56** | | | | | | | |
| Cohort 2 (OPV) | nOPV1 | 40 | 0·00 | 0·000, 0·000 | 0·00 | 0·000, 0·000 | 0·833 |
|  | mOPV1 | 20 | 0·00 | 0·000, 0·000 |  |  |  |
| Cohort 4 (OPV) | nOPV3 | 28 | 0·00 | 0·000, 0·000 | 0·00 | 0·000, 0·000 | 0·227 |
|  | mOPV3 | 15 | 0·00 | 0·000, 0·000 |  |  |  |
| N = Number of subjects in the safety population. 95% CI = Bootstrap-based 95% confidence interval for the median and difference in medians (10,000 samples). P-Value = Wilcoxon 2-sample test of a difference between groups. Log10 CCID50 per gram ≤LLOQ contributed a value equal to the LLOQ. | | | | | | | |

### **S. Table 6:** Summary of Solicited Adverse Events by Cohort, Group and Severity

|  | | **Participants** | **Severe** | **Moderate** | **Mild** | **None** |
| --- | --- | --- | --- | --- | --- | --- |
| Reaction | Vaccine | N | n (%) | n (%) | n (%) | n (%) |
| **Cohort 1** | | | | | | |
| Any Solicited Event | nOPV1 | 20 | 1 (5.0) | 3 (15.0) | 7 (35.0) | 9 (45.0) |
|  | mOPV1 | 20 | 0 (0.0) | 4 (20.0) | 8 (40.0) | 8 (40.0) |
| Fever | nOPV1 | 20 | 0 (0.0) | 0 (0.0) | 0 (0.0) | 20 (100) |
|  | mOPV1 | 20 | 0 (0.0) | 0 (0.0) | 0 (0.0) | 20 (100) |
| Chills | nOPV1 | 20 | 0 (0.0) | 0 (0.0) | 1 (5.0) | 19 (95.0) |
|  | mOPV1 | 20 | 0 (0.0) | 2 (10.0) | 3 (15.0) | 15 (75.0) |
| Fatigue | nOPV1 | 20 | 1 (5.0) | 2 (10.0) | 2 (10.0) | 15 (75.0) |
|  | mOPV1 | 20 | 0 (0.0) | 4 (20.0) | 4 (20.0) | 12 (60.0) |
| Headache | nOPV1 | 20 | 0 (0.0) | 0 (0.0) | 5 (25.0) | 15 (75.0) |
|  | mOPV1 | 20 | 0 (0.0) | 2 (10.0) | 5 (25.0) | 13 (65.0) |
| Muscle Aches/Myalgias | nOPV1 | 20 | 0 (0.0) | 1 (5.0) | 1 (5.0) | 18 (90.0) |
|  | mOPV1 | 20 | 0 (0.0) | 1 (5.0) | 5 (25.0) | 14 (70.0) |
| Joint Aches/Arthralgias | nOPV1 | 20 | 0 (0.0) | 1 (5.0) | 0 (0.0) | 19 (95.0) |
|  | mOPV1 | 20 | 0 (0.0) | 0 (0.0) | 4 (20.0) | 16 (80.0) |
| Nausea | nOPV1 | 20 | 0 (0.0) | 0 (0.0) | 2 (10.0) | 18 (90.0) |
|  | mOPV1 | 20 | 0 (0.0) | 2 (10.0) | 0 (0.0) | 18 (90.0) |
| Vomiting | nOPV1 | 20 | 0 (0.0) | 0 (0.0) | 0 (0.0) | 20 (100) |
|  | mOPV1 | 20 | 0 (0.0) | 2 (10.0) | 0 (0.0) | 18 (90.0) |
| Abdominal pain | nOPV1 | 20 | 0 (0.0) | 1 (5.0) | 4 (20.0) | 15 (75.0) |
|  | mOPV1 | 20 | 0 (0.0) | 0 (0.0) | 5 (25.0) | 15 (75.0) |
| Diarrhea | nOPV1 | 20 | 0 (0.0) | 0 (0.0) | 2 (10.0) | 18 (90.0) |
|  | mOPV1 | 20 | 0 (0.0) | 0 (0.0) | 4 (20.0) | 16 (80.0) |
| **Cohort 2: Post-Any Dose** | | | | | | |
| Any Solicited Event | nOPV1 | 50 | 1 (2.0) | 7 (14.0) | 21 (42.0) | 21 (42.0) |
|  | mOPV1 | 25 | 1 (4.0) | 7 (28.0) | 11 (44.0) | 6 (24.0) |
| Fever | nOPV1 | 50 | 0 (0.0) | 0 (0.0) | 1 (2.0) | 49 (98.0) |
|  | mOPV1 | 25 | 0 (0.0) | 0 (0.0) | 0 (0.0) | 25 (100) |
| Chills | nOPV1 | 50 | 0 (0.0) | 0 (0.0) | 2 (4.0) | 48 (96.0) |
|  | mOPV1 | 25 | 0 (0.0) | 1 (4.0) | 2 (8.0) | 22 (88.0) |
| Fatigue | nOPV1 | 50 | 0 (0.0) | 4 (8.0) | 16 (32.0) | 30 (60.0) |
|  | mOPV1 | 25 | 0 (0.0) | 4 (16.0) | 7 (28.0) | 14 (56.0) |
| Headache | nOPV1 | 50 | 0 (0.0) | 2 (4.0) | 13 (26.0) | 35 (70.0) |
|  | mOPV1 | 25 | 0 (0.0) | 4 (16.0) | 6 (24.0) | 15 (60.0) |
| Muscle Aches/Myalgias | nOPV1 | 50 | 0 (0.0) | 1 (2.0) | 8 (16.0) | 41 (82.0) |
|  | mOPV1 | 25 | 0 (0.0) | 3 (12.0) | 2 (8.0) | 20 (80.0) |
| Joint Aches/Arthralgias | nOPV1 | 50 | 0 (0.0) | 0 (0.0) | 3 (6.0) | 47 (94.0) |
|  | mOPV1 | 25 | 0 (0.0) | 1 (4.0) | 5 (20.0) | 19 (76.0) |
| Nausea | nOPV1 | 50 | 0 (0.0) | 0 (0.0) | 8 (16.0) | 42 (84.0) |
|  | mOPV1 | 25 | 1 (4.0) | 1 (4.0) | 1 (4.0) | 22 (88.0) |
| Vomiting | nOPV1 | 50 | 0 (0.0) | 0 (0.0) | 0 (0.0) | 50 (100) |
|  | mOPV1 | 25 | 1 (4.0) | 0 (0.0) | 1 (4.0) | 23 (92.0) |
| Abdominal pain | nOPV1 | 50 | 1 (2.0) | 1 (2.0) | 4 (8.0) | 44 (88.0) |
|  | mOPV1 | 25 | 0 (0.0) | 0 (0.0) | 3 (12.0) | 22 (88.0) |
| Diarrhea | nOPV1 | 50 | 0 (0.0) | 1 (2.0) | 10 (20.0) | 39 (78.0) |
|  | mOPV1 | 25 | 0 (0.0) | 1 (4.0) | 7 (28.0) | 17 (68.0) |
| **Cohort 3** | | | | | | |
| Any Solicited Event | nOPV3 | 19 | 1 (5.3) | 4 (21.1) | 5 (26.3) | 9 (47.4) |
|  | mOPV3 | 17 | 0 (0.0) | 2 (11.8) | 7 (41.2) | 8 (47.1) |
| Fever | nOPV3 | 19 | 0 (0.0) | 0 (0.0) | 0 (0.0) | 19 (100) |
|  | mOPV3 | 17 | 0 (0.0) | 0 (0.0) | 0 (0.0) | 17 (100) |
| Chills | nOPV3 | 19 | 0 (0.0) | 0 (0.0) | 1 (5.3) | 18 (94.7) |
|  | mOPV3 | 17 | 0 (0.0) | 0 (0.0) | 0 (0.0) | 17 (100) |
| Fatigue | nOPV3 | 19 | 1 (5.3) | 1 (5.3) | 5 (26.3) | 12 (63.2) |
|  | mOPV3 | 17 | 0 (0.0) | 1 (5.9) | 6 (35.3) | 10 (58.8) |
| Headache | nOPV3 | 19 | 0 (0.0) | 2 (10.5) | 1 (5.3) | 16 (84.2) |
|  | mOPV3 | 17 | 0 (0.0) | 0 (0.0) | 5 (29.4) | 12 (70.6) |
| Muscle Aches/Myalgias | nOPV3 | 19 | 0 (0.0) | 0 (0.0) | 2 (10.5) | 17 (89.5) |
|  | mOPV3 | 17 | 0 (0.0) | 1 (5.9) | 1 (5.9) | 15 (88.2) |
| Joint Aches/Arthralgias | nOPV3 | 19 | 0 (0.0) | 0 (0.0) | 1 (5.3) | 18 (94.7) |
|  | mOPV3 | 17 | 0 (0.0) | 0 (0.0) | 2 (11.8) | 15 (88.2) |
| Nausea | nOPV3 | 19 | 0 (0.0) | 0 (0.0) | 1 (5.3) | 18 (94.7) |
|  | mOPV3 | 17 | 0 (0.0) | 0 (0.0) | 1 (5.9) | 16 (94.1) |
| Vomiting | nOPV3 | 19 | 0 (0.0) | 0 (0.0) | 1 (5.3) | 18 (94.7) |
|  | mOPV3 | 17 | 0 (0.0) | 0 (0.0) | 0 (0.0) | 17 (100) |
| Abdominal pain | nOPV3 | 19 | 0 (0.0) | 3 (15.8) | 3 (15.8) | 13 (68.4) |
|  | mOPV3 | 17 | 0 (0.0) | 0 (0.0) | 1 (5.9) | 16 (94.1) |
| Diarrhea | nOPV3 | 19 | 0 (0.0) | 0 (0.0) | 3 (15.8) | 16 (84.2) |
|  | mOPV3 | 17 | 0 (0.0) | 1 (5.9) | 5 (29.4) | 11 (64.7) |
| **Cohort 4: Post-Any Dose** | | | | | | |
| Any Solicited Event | nOPV3 | 35 | 1 (2.9) | 7 (20.0) | 7 (20.0) | 20 (57.1) |
|  | mOPV3 | 19 | 1 (5.3) | 3 (15.8) | 6 (31.6) | 9 (47.4) |
| Fever | nOPV3 | 35 | 0 (0.0) | 0 (0.0) | 0 (0.0) | 35 (100) |
|  | mOPV3 | 19 | 0 (0.0) | 0 (0.0) | 1 (5.3) | 18 (94.7) |
| Chills | nOPV3 | 35 | 0 (0.0) | 0 (0.0) | 1 (2.9) | 34 (97.1) |
|  | mOPV3 | 19 | 0 (0.0) | 0 (0.0) | 1 (5.3) | 18 (94.7) |
| Fatigue | nOPV3 | 35 | 1 (2.9) | 5 (14.3) | 3 (8.6) | 26 (74.3) |
|  | mOPV3 | 19 | 1 (5.3) | 2 (10.5) | 4 (21.1) | 12 (63.2) |
| Headache | nOPV3 | 35 | 0 (0.0) | 2 (5.7) | 6 (17.1) | 27 (77.1) |
|  | mOPV3 | 19 | 0 (0.0) | 3 (15.8) | 2 (10.5) | 14 (73.7) |
| Muscle Aches/Myalgias | nOPV3 | 35 | 0 (0.0) | 0 (0.0) | 2 (5.7) | 33 (94.3) |
|  | mOPV3 | 19 | 0 (0.0) | 1 (5.3) | 1 (5.3) | 17 (89.5) |
| Joint Aches/Arthralgias | nOPV3 | 35 | 0 (0.0) | 0 (0.0) | 2 (5.7) | 33 (94.3) |
|  | mOPV3 | 19 | 0 (0.0) | 1 (5.3) | 0 (0.0) | 18 (94.7) |
| Nausea | nOPV3 | 35 | 0 (0.0) | 2 (5.7) | 6 (17.1) | 27 (77.1) |
|  | mOPV3 | 19 | 0 (0.0) | 0 (0.0) | 1 (5.3) | 18 (94.7) |
| Vomiting | nOPV3 | 35 | 0 (0.0) | 0 (0.0) | 1 (2.9) | 34 (97.1) |
|  | mOPV3 | 19 | 0 (0.0) | 0 (0.0) | 0 (0.0) | 19 (100) |
| Abdominal pain | nOPV3 | 35 | 0 (0.0) | 1 (2.9) | 4 (11.4) | 30 (85.7) |
|  | mOPV3 | 19 | 0 (0.0) | 0 (0.0) | 2 (10.5) | 17 (89.5) |
| Diarrhea | nOPV3 | 35 | 0 (0.0) | 1 (2.9) | 4 (11.4) | 30 (85.7) |
|  | mOPV3 | 19 | 0 (0.0) | 1 (5.3) | 4 (21.1) | 14 (73.7) |
| N= Number of Subjects in the safety population who received the specified dose. n = number of subjects with at least one event. Cells display number (%) of subjects with at least one event at the specified severity. The maximum severity reported is used for summaries across events (Any Solicited Event). | | | | | | |
